# Mpox coinfections and clinical manifestation in Africa: a systematic review and meta-analysis

**DOI:** 10.64898/2026.02.06.26345747

**Authors:** Fabrice Zobel Lekeumo Cheuyem, Chabeja Achangwa, Lionel Berthold Keubou Boukeng, Rick Tchamani, Jessica Davies, Christian Noël Malaka, Armel Evouna Mbarga

## Abstract

**Background:** Africa continues to have a significant public health problem with mpox, where endemic transmission persists and overlaps with a high burden of other infectious diseases. Although their epidemiology and clinical impact are still poorly understood throughout the continent, coinfections with human immunodeficiency virus (HIV) and varicella-zoster virus (VZV) can affect the clinical picture, severity of the disease, and accuracy of the diagnosis.

**Methods:** Following PRISMA criteria, we registered the protocol in PROSPERO (CRD420251133960) and carried out a methodical review and meta-analysis. Through searches of numerous electronic databases and grey literature up to February 27, 2025, observational studies documenting mpox coinfections with VZV and/or HIV and related clinical symptoms in Africa were searched. Pooled prevalence was calculated using random-effects models. Subgroup analyses and meta-regression were conducted to investigate heterogeneity across WHO regions, countries, study designs, settings and type of participants.

**Results:** A total of 27 studies carried out across African countries were included. The pooled prevalence of VZV–mpox coinfection was 10.23% (95% CI: 2.95–29.93), while HIV–mpox coinfection prevalence was 6.55% (95% CI: 2.36–16.90), both of which had significant heterogeneity. Coinfections were far more prevalent in hospital-based environments than in community-based research. The rash was nearly universal throughout all clades, but the clinical manifestations varied depending on the viral clade, with clades I and Ia linked to more severe systemic symptoms than clade II.

**Discussions:** HIV and VZV coinfections with mpox pose a major yet possibly underestimated burden in Africa and are linked to more severe clinical presentations, particularly in hospital environments. The necessity of including clinical, epidemiological, and genomic data into mpox monitoring systems is emphasized by the observed clinical differences between clades. Improving patient management and outbreak preparedness across the continent requires strengthening diagnostic capacity and routinely screening for coinfections.

## 1. Introduction

Mpox is a re-emerging zoonotic viral infection, caused by a stable DNA *Orthopoxvirus* belonging to the *Poxviridae* family, that has now become a significant global health concern (Sah et al., 2022; Africa CDC Declares Mpox A Public Health Emergency of Continental Security, Mobilizing Resources Across the Continent, 2024). During the 2022–2023 multicountry outbreak, more than 87,000 confirmed cases were reported globally, with the United States and Brazil ranking among the most affected countries, underscoring its potential for rapid international spread (Di Gennaro et al., 2022; Multi-country monkeypox outbreak: situation update, 2022). In Africa, the epidemiological burden remains disproportionately high (Cheuyem et al., 2025b). In Central Africa, the Democratic Republic of the Congo (DRC) reported over 14,600 suspected cases and >500 deaths between 2023 and 2024, and Cameroon continues to register recurrent clusters across multiple regions (Bangwen et al., 2025; Brosius et al., 2025; Ebede et al., 2025). In West Africa, Nigeria has documented more than 1,400 suspected and confirmed cases since 2017, with resurgences in 2022 and 2023, while Ghana recorded confirmed outbreaks in 2022, including pediatric cases (Cadmus et al., 2024; WHO Director-General declares mpox outbreak a public health emergency of international concern, 2024; Cibenda et al., 2025). Coinfections with endemic pathogens particularly HIV, bacterial skin infections, sexually transmitted infections, and varicella-zoster virus (VZV) are widespread in these settings and may significantly modulate disease severity, transmission dynamics, and clinical outcomes (Hughes et al., 2021; Cheuyem et al., 2025a).

Mpox virus is classified into two main clades. Clade I, historically associated with Central Africa, is linked to higher virulence and has recently been subdivided into Ia and Ib, with Clade Ib emerging in the DRC in 2023 and demonstrating greater transmissibility (Iroezindu et al., 2023; Brosius et al., 2025). Clade II, predominant in West Africa, includes Clade IIb, which drove the global outbreak between 2022 and 2023 and is associated with milder clinical outcomes (Multi-country monkeypox outbreak: situation update, 2022; Iroezindu et al., 2023). Classically, mpox presents with a febrile prodrome followed by a vesiculopustular rash and lymphadenopathy. However, clinical manifestations in Africa are often modified by co-existing infections (Hughes et al., 2021; Cheuyem et al., 2025a). These coinfections may significantly modulate disease severity, transmission dynamics, and outcomes. In resource-constrained settings, the overlapping burden of mpox and these infectious diseases presents unique challenges for clinical management and public health control, making research into coinfection dynamics both urgent and essential (Hughes et al., 2021; Cheuyem et al., 2025a). Accumulating evidence shows that coinfected patients may exhibit atypical rashes, prolonged illness, or more severe systemic manifestations, although most available data arise from small studies, outbreak investigations, or case reports.

Despite growing documentation, significant uncertainties persist regarding mpox coinfections in Africa. There is no comprehensive estimate of coinfection prevalence, partly due to fragmented surveillance systems and limited laboratory capacity (Hughes et al., 2021; Cheuyem et al., 2025a). Moreover, the clinical impact of specific coinfections, especially HIV, VZV, and bacterial infections, remains poorly quantified. It is unclear how coinfections modify rash morphology, duration of viral shedding, immune response, or risk of complications (Hughes et al., 2021; Cheuyem et al., 2025a). Another unresolved gap concerns interactions between viral clades, coinfection profiles and clinical manifestation (Iroezindu et al., 2023; Brosius et al., 2025). Additionally, mpox shares clinical similarities with several endemic infections, making misdiagnosis common. Few studies have systematically assessed diagnostic overlap (Hughes et al., 2021; Cheuyem et al., 2025a). Finally, the lack of large multicountry cohort data limits our understanding of risk factors for severe disease among coinfected individuals and constrains the development of evidence-based clinical guidelines.

Addressing these gaps is vital for both clinical practice and public health. Improved knowledge of coinfection prevalence and clinical impact would support early risk stratification, refine case definitions, and inform treatment protocols tailored to high-burden settings (Hughes et al., 2021; Cheuyem et al., 2025a). From a public-health perspective, understanding how coinfections interact with viral clades can strengthen surveillance systems, guide vaccination and testing strategies, and optimize resource allocation in low-resource contexts (Iroezindu et al., 2023; Cheuyem et al., 2025a). Such evidence is necessary to prevent severe outcomes, improve outbreak preparedness, and ensure effective and equitable control of mpox across Africa. This systematic review therefore aims to describe the landscape of coinfections (VZV and HIV) among confirmed mpox cases and characterize the trend of clinical presentation of clade categorized mpox patients in Africa.

## 2. Methods

### 2.1. Study design

The Preferred Reporting Items for Systematic Review and Meta-analysis (PRISMA) guidelines was applied while reporting the study (Moher et al., 2009). To ensure process rigor and transparency, the study protocol was filed in the International Prospective Register of Systematic Reviews (CRD420251133960).

### 2.2. Eligibility criteria

Any observational studies that report on patients with mpox coinfections and clinical manifestation, conducted in Africa were included. Populations of interest included both community-based cases and hospital-based cases, including adults, children. The key exposures of interest were coinfections occurring in mpox patients (HIV, VZV). To capture the full historical and contemporary evidence, the timeframe spanned till 2024. Only studies published in English or French were considered.

### 2.3. Exclusion criteria

Studies were excluded for the following reasons: duplication of data, focus outside the scope of our research objectives, non-observational studies (comment, letter to editor, review, other systematic review) and studies conducted outside Africa. We also excluded studies focusing exclusively on laboratory, genomic, or animal models without reporting clinical or coinfection data. Additionally, articles lacking full-text availability were omitted due to insufficient data or absence of required outcome measures.

### 2.4. Article searching strategy

A comprehensive search strategy was employed to identify all relevant literature. Multiple electronic databases were searched, including PubMed, Scopus, ScienceDirect, Web of Science, CINAHL, and EMBASE. To incorporate research from African scholars, African Journals Online (AJOL) was also consulted. In addition to these databases, we searched for grey literature, which includes unpublished research and preprints. Moreover, manual search was conducted on google scholar and on reference lists of included studies. For PubMed the search strategy was a combination of search MeSH terms: (“Monkeypox” OR “Mpox” OR “Monkeypox virus” OR “MPXV”, “Varicella-zoster virus’ OR “HIV,”), AND (“Africa,” OR individual country names such as “Democratic Republic of Congo,” “Nigeria,” or “Cameroon”…). Additionally, a manual search for additional publications that were not indexed in these databases. The final search was concluded on February 27, 2025.

### 2.5. Data extraction

We developed a Microsoft Excel 2016 form to collect study characteristics from all included study reports. This form captured the first author’s name, study year, region, study design, type of participant, setting, sampling method, total number of confirmed mpox cases, the total number VZV cases, number of VZV and mpox coinfected patients, the number of HIV and mpox coinfected patients et total number of patients tested for VZV, clade categorization, and frequency of each clinical manifestation. One investigator (FZLC) extracted to data and two additional investigators (CA and AEM) independently verified the extracted data for accuracy.

### 2.6. Data quality assessment

The Joanna Briggs Institute (JBI) quality assessment tool was used to evaluate the quality of studies included (JBI Critical Appraisal Tools, 2017). This was conducted by two independent reviewers (FZLC and LBKB). Risk of bias was assessed using nine or ten criteria, depending on the study design. (1) For cross-sectional studies, criteria included: appropriateness of the sampling frame, use of a suitable sampling technique, adequate sample size, description of study subjects and setting, sufficient data analysis, use of valid methods for identifying conditions and measurements, use of appropriate statistical analysis, and an adequate response rate (≥60%). (2) For case series, criteria included: standardized measurement and valid identification of the condition for all participants, consecutive and complete inclusion of participants, reporting of participant demographics and clinical information, reporting of outcomes or follow-up results, reporting of the presenting site(s)/clinic(s) demographic information, and statistical analysis appropriate for case series studies. (3) For case reports, criteria included: clear description of patient demographic characteristics, patient history and timeline, current clinical condition on presentation, diagnostic tests/assessment methods and results, interventions/treatment procedures, post-intervention clinical condition, adverse/unanticipated events, and takeaway lessons. Each criterion was scored as 1 (yes) or 0 (no or unclear). The overall risk of bias was categorized as low (>50%), moderate (>25-50%), or high (≤25%). Any disagreements between the reviewers were resolved through discussion or by consultation with a third reviewer (AEM).

### 2.7. Outcome measurement

The primary outcomes of this systematic review and meta-analysis were the coinfection rate of mpox and VZV, or HIV. Secondary outcomes included The VZV prevalence and mpox clinical manifestation by clade classification. Mpox coinfection rate was calculated by dividing the number of confirmed VZV or HIV cases by the total number of confirmed Mpox cases. The prevalence of VZV was calculated by dividing the number of confirmed VZV cases by the total number of patients tested. For clinical features among confirmed Mpox cases, the proportion of each manifestation was determined by dividing its frequency by the number of clinically assessed confirmed mpox cases.

### 2.8. Operational definition

A patient was considered laboratory-confirmed Mpox if at least one specimen tested positive for Orthopoxvirus using a specific assay or Mpox-specific real-time PCR, or if Mpox was isolated in culture. A case was defined as laboratory-confirmed VZV if at least one specimen yielded a positive result in a real-time PCR assay targeting the VZV-specific DNA signature (Whitehouse et al., 2021). Similarly, a case was defined as laboratory-confirmed HIV if at least one specimen showed a positive result in an HIV antigen-specific assay or HIV-specific real-time PCR (Hutin et al., 2001; Brosius et al., 2025).

### 2.9. Statistical analysis and synthesis

Study heterogeneity was assessed using the Cochrane Q statistic and the heterogeneity between studies was evaluated using the I² statistic, which classified it as low (<25%), moderate (25-75%), or high (>75%). A random-effects model was applied for all pooled analyses. To investigate potential sources of heterogeneity, we performed subgroup analyses based on study period, country, setting, participant type. Countries were categorized according to the WHO African Region classification (ESPEN, 2023): Western (Nigeria), Eastern (Burundi, Kenya), and Central (Cameroon, Central African Republic, The DRC). The association between these study characteristics and the pooled estimates was further examined using univariable and multivariable meta-regression. We utilized the Generalized Linear Mixed Models (GLMM), coupled with the Probit-Logit Transformation (PLOGIT), which is robust for synthesizing proportional data, including extreme proportions of 0% or 100%, without needing continuity corrections (Stijnen et al., 2010). Statistical significance was defined as a two-sided *p*-value < 0.05. All analyses were performed using R software version 4.5.1 with the ‘meta’ package (R Core Team, 2024).

#### Publication bias and sensitivity test

Publication bias was assessed visually using a funnel plot. The asymmetry of the inverted funnel shape suggested the potential of publication bias. In addition, statistical evaluation was conducted using Egger’s linear regression test and Begg’s rank correlation test, with a *p*-value < 0.05 indicating significant risk of publication bias. The trim-and-fill method was used to adjust for potential missing studies (Shi and Lin, 2019). To test the robustness of the findings we performed sensitivity analysis by iteratively excluding one study at a time and pooled the resulting estimate.

## 3. Results

A total of 3,779 records were obtained from database searches, along with three additional records from other sources. After removing 448 duplicates, 3,334 articles were screened based on their titles and abstracts, resulting in the exclusion of 3,157 for not satisfying our inclusion criteria. The full texts of 177 articles were then evaluated for eligibility. An additional 150 articles were excluded because they did not report the outcomes of interest (n = 148) or were review articles (n = 2). In the end, 27 studies met all criteria and were included in the systematic review and meta-analysis (Fig. 1).

**Fig. 1.**
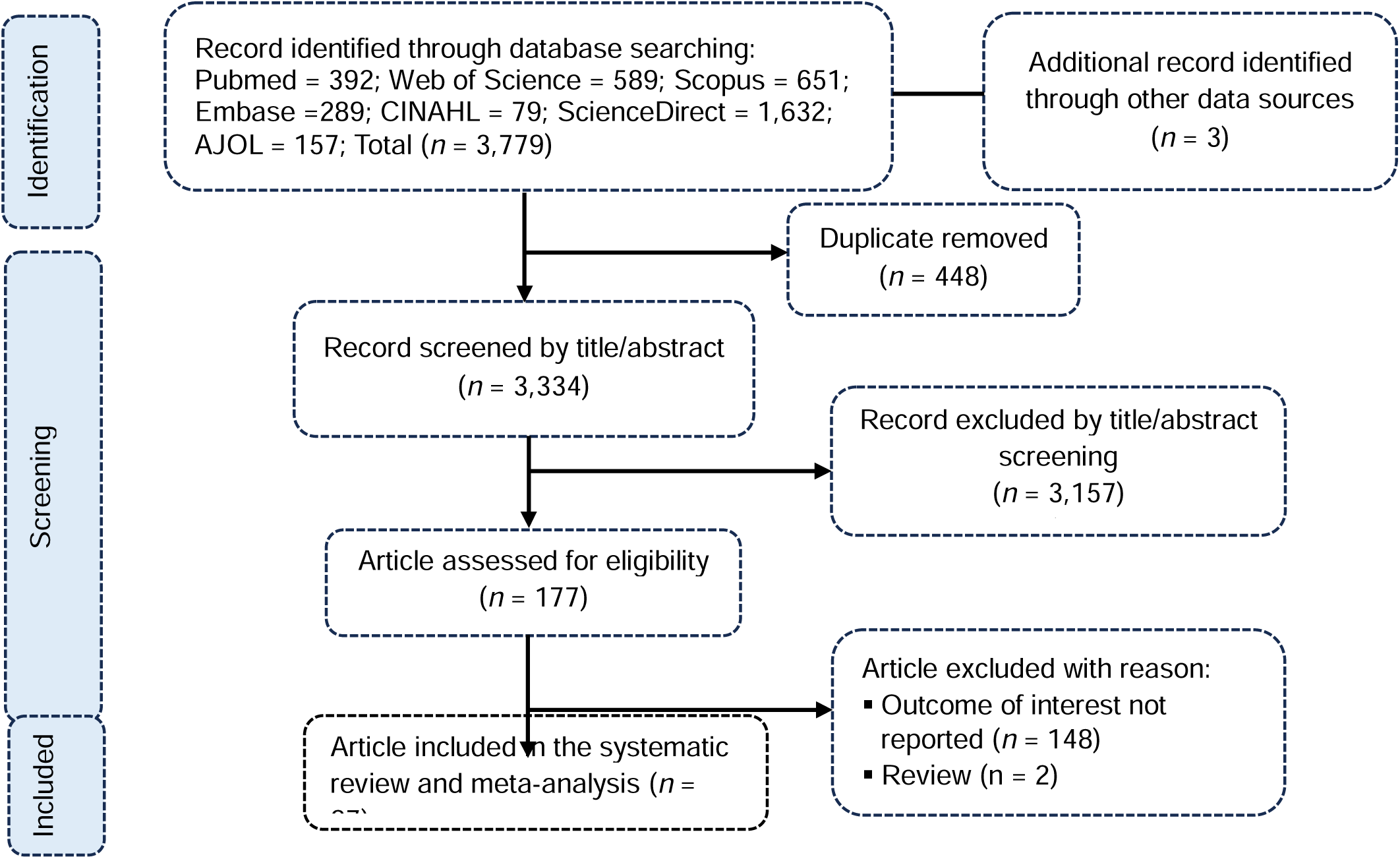
PRISMA diagram flow from study identification to inclusion in the meta-analysis

### 3.1. Studies selection

### 3.2. Characteristics of studies included

A total of 27 studies from the Democratic Republic of the Congo (DRC; 59.3%; n = 16 studies) (Mwanba et al., 1997; Hutin et al., 2001; Meyer et al., 2002; McCollum et al., 2015; Nolen et al., 2016; Hoff et al., 2017; Osadebe et al., 2017; Petersen et al., 2019; Hughes et al., 2021; Whitehouse et al., 2021; Mande et al., 2022; Kinganda-Lusamaki et al., 2023; Masirika et al., 2024; Vakaniaki et al., 2024; Bangwen et al., 2025; Brosius et al., 2025), Nigeria (25.9%; n = 7 studies) (Ogoina et al., 2019, 2020; Echekwube et al., 2020; Ogoina and Yinka-Ogunleye, 2022; Stephen et al., 2022; Mmerem et al., 2024a, 2024b), the Central African Republic (CAR; 3.7%; n = 1 study) (Besombes et al., 2023), Cameroon (3.7%; n = 1 study) (Djuicy et al., 2024), Kenya (3.7%; n = 1 study) (Onyango et al., 2025) and Burundi (3.7%; n = 1 study) (Nizigiyimana et al., 2024) were included. Most were cross-sectional (81.5%; n = 22 studies), community-based (55.6%; n = 15 studies) with low risk of bias (96.3%; n = 26 studies). Coinfection rates of VZV and mpox (reported in 13 studies) varied widely, from 0% in some studies to 81-100% in others. All included studies used-non-probabilistic sampling (Table 1).

**Table 1.**
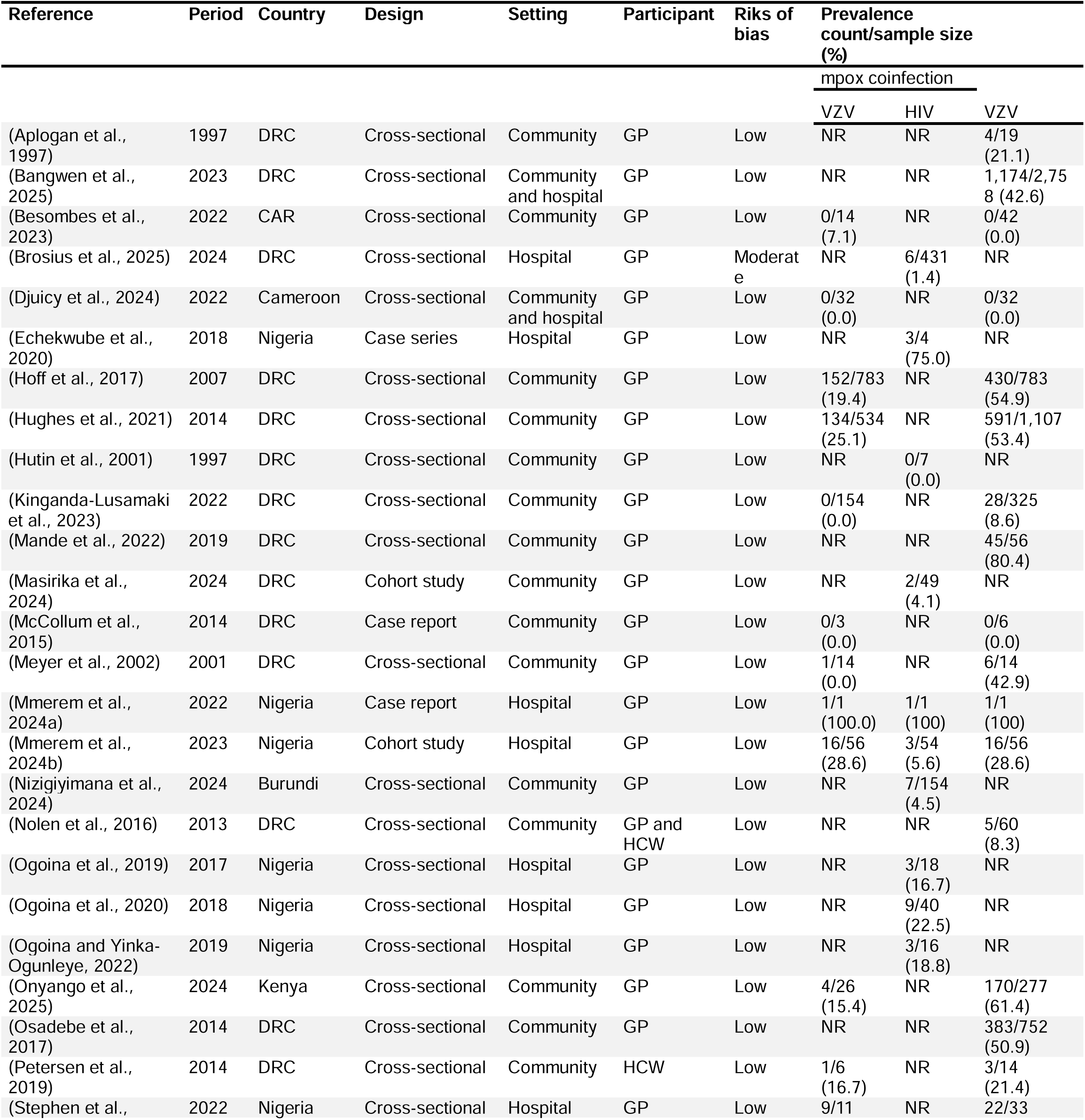
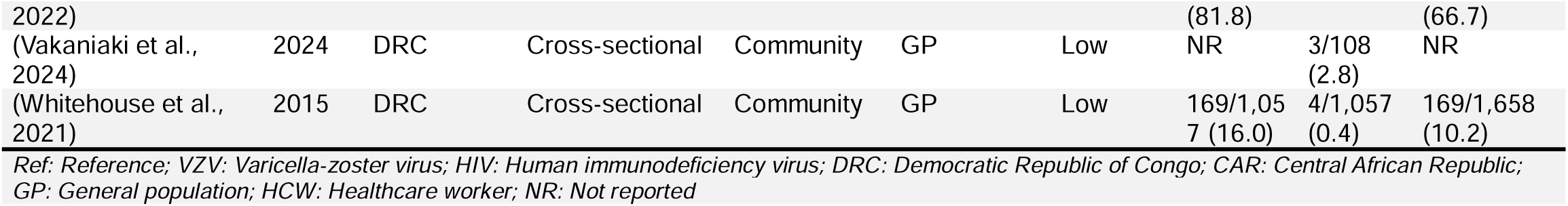
Characteristics of included studies.

### 3.3. VZV coinfection among confirmed mpox cases in Africa

Across 13 studies (Meyer et al., 2002; McCollum et al., 2015; Hoff et al., 2017; Petersen et al., 2019; Hughes et al., 2021; Whitehouse et al., 2021; Stephen et al., 2022; Besombes et al., 2023; Kinganda-Lusamaki et al., 2023; Djuicy et al., 2024; Mmerem et al., 2024a, 2024b; Onyango et al., 2025) (n = 2,691), the pooled prevalence of VZV–mpox coinfection was 10.23% (95% CI: 2.95–29.93). Heterogeneity was (*I*² = 67.7%, *p* ˂0.001), indicating moderate heterogeneity between-study differences and suggesting that the pooled estimate should be viewed with caution across different settings, populations and other methodological characteristics (Fig. 2).

**Fig. 2.**
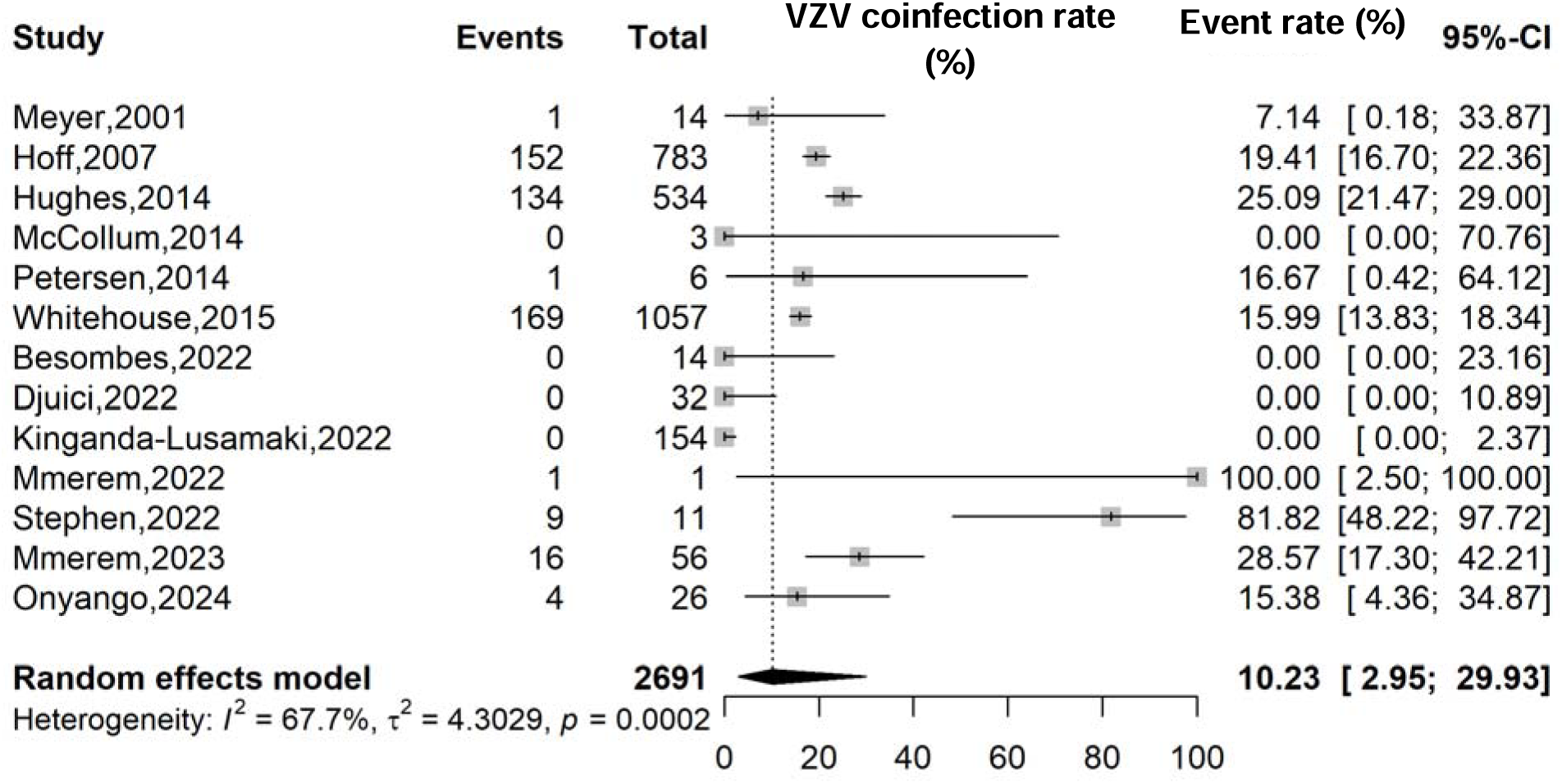
Varicella-zoster virus (VZV) infection prevalence among confirmed mpox cases in Africa

Subgroup analyses showed a higher pooled prevalence of VZV–mpox coinfection in studies conducted before 2022 (19.14%; 95% CI: 15.43–23.51; 2,397 participants; n = 6 studies) compared to those completed from 2022 onward (5.54%; 95% CI: 0.17–66.48; 294 participants; n = 7 studies), though this difference was not statistically significant (*p* = 0.438). A strong and statistically significant difference was observed based on study setting (*p* < 0.001), with hospital-based studies showing much higher prevalence (60.66%; 95% CI: 19.46–90.78; 68 participants, n = 3 studies) than community-based studies (7.60%; 95% CI: 2.51–20.79; 2,591 participants; n = 9 studies). Although differences between countries were not statistically significant (*p* = 0.187), the highest pooled prevalence was observed in Nigeria (60.66%; 95% CI: 19.46–90.78; 68 participants; n = 3 studies), followed by Kenya (15.38%; 95% CI: 4.36–34.87; 26 participants, n = 1 study) and the DRC (8.04%; 95% CI: 2.25–24.92; 2,551 participants, n = 7 studies). WHO regional analysis showed a significant gradient (*p* < 0.001), with the highest prevalence in Western Africa (60.66%; 95% CI: 19.46–90.78; 68 participants, n = 3 studies) and the lowest in Central Africa (4.28%; 95% CI: 0.87–18.51; 2,597 participants; n = 9 studies) (Table 2).

**Table 2.**
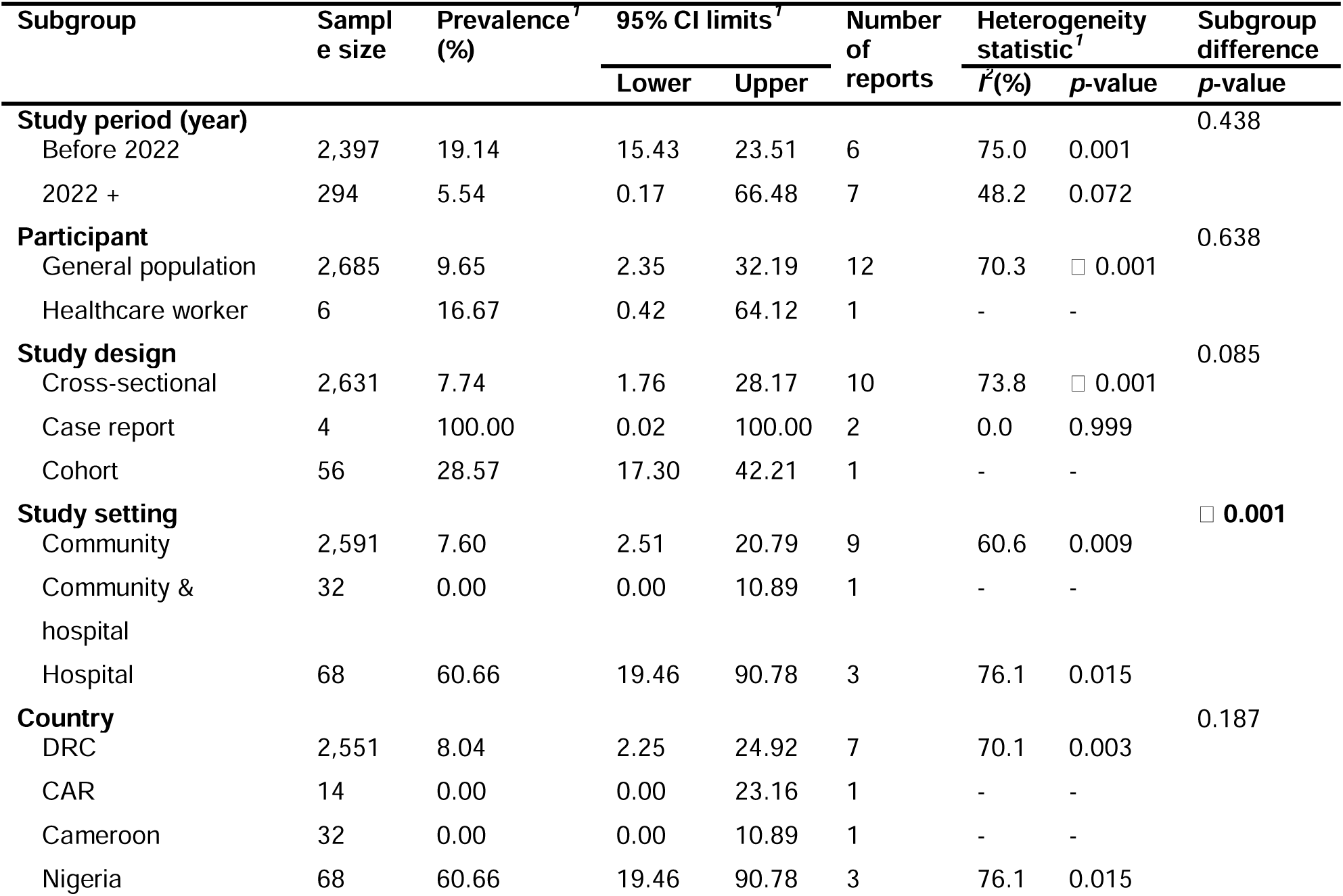
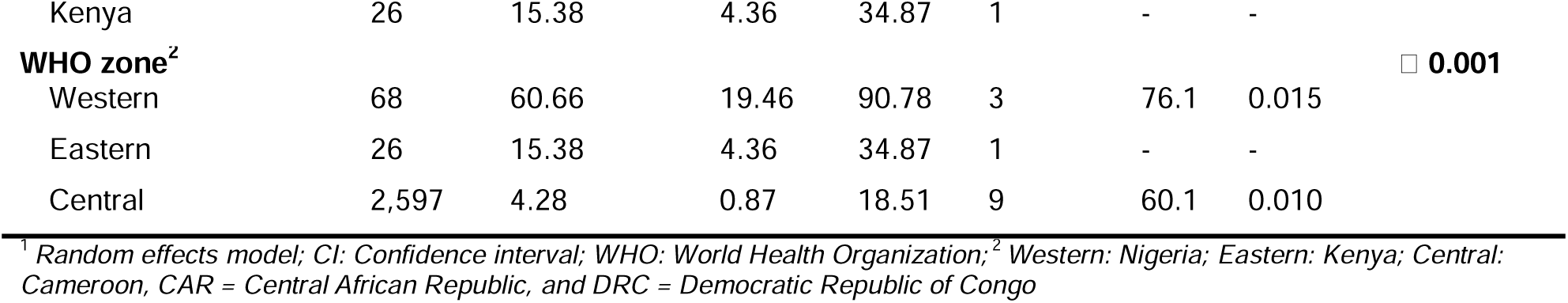
Subgroup meta-analysis of the pooled prevalence of varicella-zoster virus coinfection among confirmed mpox cases in Africa.

| Subgroup | Sample size | Prevalence<sup>†</sup> (%) | 95% CI limits<sup>†</sup> |  | Number of reports | Heterogeneity statistic<sup>†</sup> |  | Subgroup difference p-value |
| --- | --- | --- | --- | --- | --- | --- | --- | --- |
| | | | Lower | Upper | | $I^2$ (%) | p-value | |
| **Study period (year)** |  |  |  |  |  |  |  | 0.438 |
| Before 2022 | 2,397 | 19.14 | 15.43 | 23.51 | 6 | 75.0 | 0.001 |  |
| 2022 + | 294 | 5.54 | 0.17 | 66.48 | 7 | 48.2 | 0.072 |  |
| **Participant** |  |  |  |  |  |  |  | 0.638 |
| General population | 2,685 | 9.65 | 2.35 | 32.19 | 12 | 70.3 | □ 0.001 |  |
| Healthcare worker | 6 | 16.67 | 0.42 | 64.12 | 1 | - | - |  |
| **Study design** |  |  |  |  |  |  |  | 0.085 |
| Cross-sectional | 2,631 | 7.74 | 1.76 | 28.17 | 10 | 73.8 | □ 0.001 |  |
| Case report | 4 | 100.00 | 0.02 | 100.00 | 2 | 0.0 | 0.999 |  |
| Cohort | 56 | 28.57 | 17.30 | 42.21 | 1 | - | - |  |
| **Study setting** |  |  |  |  |  |  |  | □ 0.001 |
| Community | 2,591 | 7.60 | 2.51 | 20.79 | 9 | 60.6 | 0.009 |  |
| Community & hospital | 32 | 0.00 | 0.00 | 10.89 | 1 | - | - |  |
| Hospital | 68 | 60.66 | 19.46 | 90.78 | 3 | 76.1 | 0.015 |  |
| **Country** |  |  |  |  |  |  |  | 0.187 |
| DRC | 2,551 | 8.04 | 2.25 | 24.92 | 7 | 70.1 | 0.003 |  |
| CAR | 14 | 0.00 | 0.00 | 23.16 | 1 | - | - |  |
| Cameroon | 32 | 0.00 | 0.00 | 10.89 | 1 | - | - |  |
| Nigeria | 68 | 60.66 | 19.46 | 90.78 | 3 | 76.1 | 0.015 |  |

|  |  |  |  |  |  |  |  |
| --- | --- | --- | --- | --- | --- | --- | --- |
| Kenya | 26 | 15.38 | 4.36 | 34.87 | 1 | - | - |
| **WHO zone<sup>2</sup>** |  |  |  |  |  |  | **□ 0.001** |
| Western | 68 | 60.66 | 19.46 | 90.78 | 3 | 76.1 | 0.015 |
| Eastern | 26 | 15.38 | 4.36 | 34.87 | 1 | - | - |
| Central | 2,597 | 4.28 | 0.87 | 18.51 | 9 | 60.1 | 0.010 |

<sup>1</sup> Random effects model; CI: Confidence interval; WHO: World Health Organization; <sup>2</sup> Western: Nigeria; Eastern: Kenya; Central: Cameroon, CAR = Central African Republic, and DRC = Democratic Republic of Congo

### 3.4. HIV coinfection among confirmed mpox cases in Africa

Across 12 studies (Hutin et al., 2001; Ogoina et al., 2019, 2020; Whitehouse et al., 2021; Ogoina and Yinka-Ogunleye, 2022; Masirika et al., 2024; Mmerem et al., 2024a, 2024b; Nizigiyimana et al., 2024; Vakaniaki et al., 2024; Brosius et al., 2025) involving 1,939 confirmed mpox cases, the pooled prevalence of HIV coinfection was 6.55% (95% CI: 2.36–16.90; *I*² = 86.5%, *p* < 0.0001). Heterogeneity was high, indicating substantial difference between studies and suggesting that the pooled estimate should be interpreted with caution (Fig. 3).

**Fig. 3.**
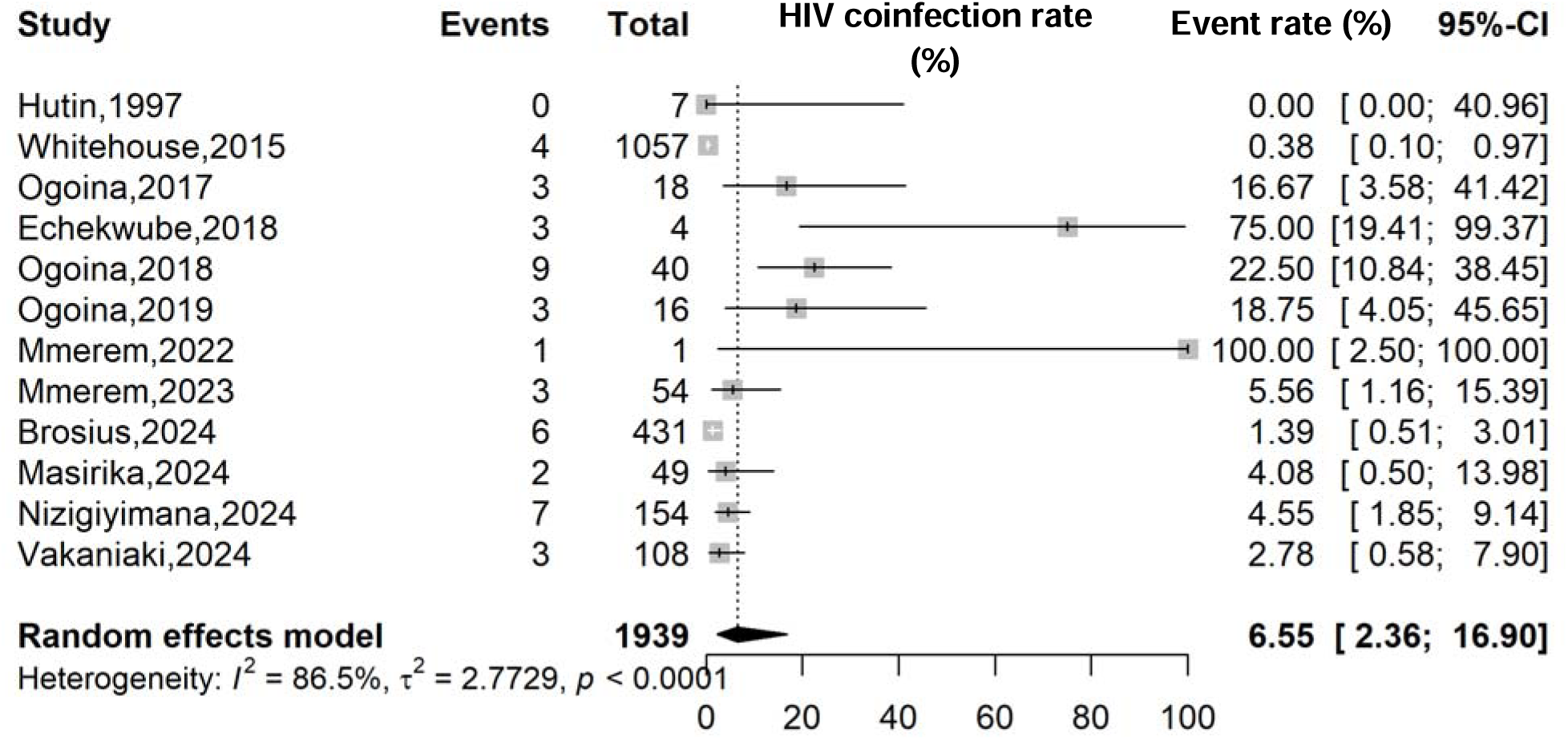
Human immunodeficiency virus (HIV) infection prevalence among confirmed mpox cases in Africa

Subgroup analyses showed lower pooled prevalence of HIV coinfection in studies published from 2022 onward (3.14%; 95% CI: 1.74–5.61; 1,142 participants, n = 6 studies) compared with studies conducted before 2022 (9.48%; 95% CI: 1.66–39.47; 797 participants, n = 6 studies), although this difference was not statistically significant (*p* = 0.233). A significant difference was observed by study setting (*p* = 0.007), with hospital-based studies reporting higher prevalence (15.97%; 95% CI: 4.67–42.44; 564 participants; n = 7 studies) than community-based studies (1.75%; 95% CI: 0.62–4.86; 1,375 participants; n = 5 studies). Country-level differences were statistically significant (*p* < 0.001), with higher prevalence of HIV coinfection in Nigeria (21.19%; 95% CI: 8.82–42.77; 133 participants; n = 6 studies) than in the DRC (1.21%; 95% CI: 0.49–2.94; 1,652 participants; n = 5 studies). WHO regional analysis mirrored country-level findings (*p* < 0.001), with the highest prevalence in the Western WHO zone (21.19%; 95% CI: 8.82–42.77; 133 participants; n = 6 studies) and the lowest in Central zone (1.21%; 95% CI: 0.49–2.94; 1,652 participants; n = 5 studies) (Table 3).

**Table 3.** Subgroup meta-analysis of the pooled prevalence of varicella-zoster virus coinfection among confirmed mpox cases in Africa.

| Subgroup | Sample size | Prevalence<sup>1</sup> (%) | 95% CI limits<sup>1</sup> |  | Number of reports | Heterogeneity statistic<sup>1</sup> |  | Subgroup difference p-value |
| --- | --- | --- | --- | --- | --- | --- | --- | --- |
| | | | Lower | Upper | | $I^2$ (%) | p-value | |
| **Study period (year)** |  |  |  |  |  |  |  | 0.233 |
| Before 2022 | 1,142 | 9.48 | 1.66 | 39.47 | 6 | 91.9 | □ 0.001 |  |
| 2022 + | 797 | 3.14 | 1.74 | 5.61 | 6 | 20.1 | 0.282 |  |
| **Study design** |  |  |  |  |  |  |  | **0.008** |
| Cross-sectional | 1,831 | 3.99 | 1.37 | 11.05 | 8 | 89.8 | □ 0.001 |  |
| Cohort | 103 | 4.85 | 2.03 | 11.14 | 2 | 0.0 | 0.729 |  |
| Case report | 1 | 100.00 | 2.50 | 100.00 | 1 | - | - |  |
| Case series | 4 | 75.00 | 19.42 | 99.37 | 1 | - | - |  |
| **Study setting** |  |  |  |  |  |  |  | **0.007** |
| Community | 1,375 | 1.75 | 0.62 | 4.86 | 5 | 76.9 | 0.002 |  |
| Hospital | 564 | 15.97 | 4.67 | 42.44 | 7 | 86.0 | □ 0.001 |  |
| **Country** |  |  |  |  |  |  |  | □ **0.001** |
| DRC | 1,652 | 1.21 | 0.49 | 2.94 | 5 | 62.2 | 0.032 |  |
| Nigeria | 133 | 21.19 | 8.82 | 42.77 | 6 | 52.9 | 0.060 |  |
| Burundi | 154 | 4.55 | 1.85 | 9.14 | 1 | - | - |  |
| **WHO zone<sup>2</sup>** |  |  |  |  |  |  |  | □ **0.001** |
| Western | 133 | 21.19 | 8.82 | 42.77 | 6 | 52.9 | 0.060 |  |
| Eastern | 154 | 4.55 | 1.85 | 9.14 | 1 | - | - |  |
| Central | 1,652 | 1.21 | 0.49 | 2.94 | 5 | 62.2 | 0.032 |  |

<sup>1</sup> Random effects model; CI: Confidence interval; WHO: World Health Organization; <sup>2</sup> Western: Nigeria; Eastern: Kenya; Central: Cameroon, CAR = Central African Republic, and DRC = Democratic Republic of Congo

### 3.5. Varicella-zoster virus infections in Africa

Across 18 studies involving 7,993 participants, the overall prevalence of varicella-zoster virus (VZV) infection in Africa was 26.35% (95% CI: 13.84–44.35; *I*² = 98.0%, *p* < 0.001). The high heterogeneity suggested that the pooled prevalence estimate should be interpreted with caution (Fig. 4).

**Fig. 4.**
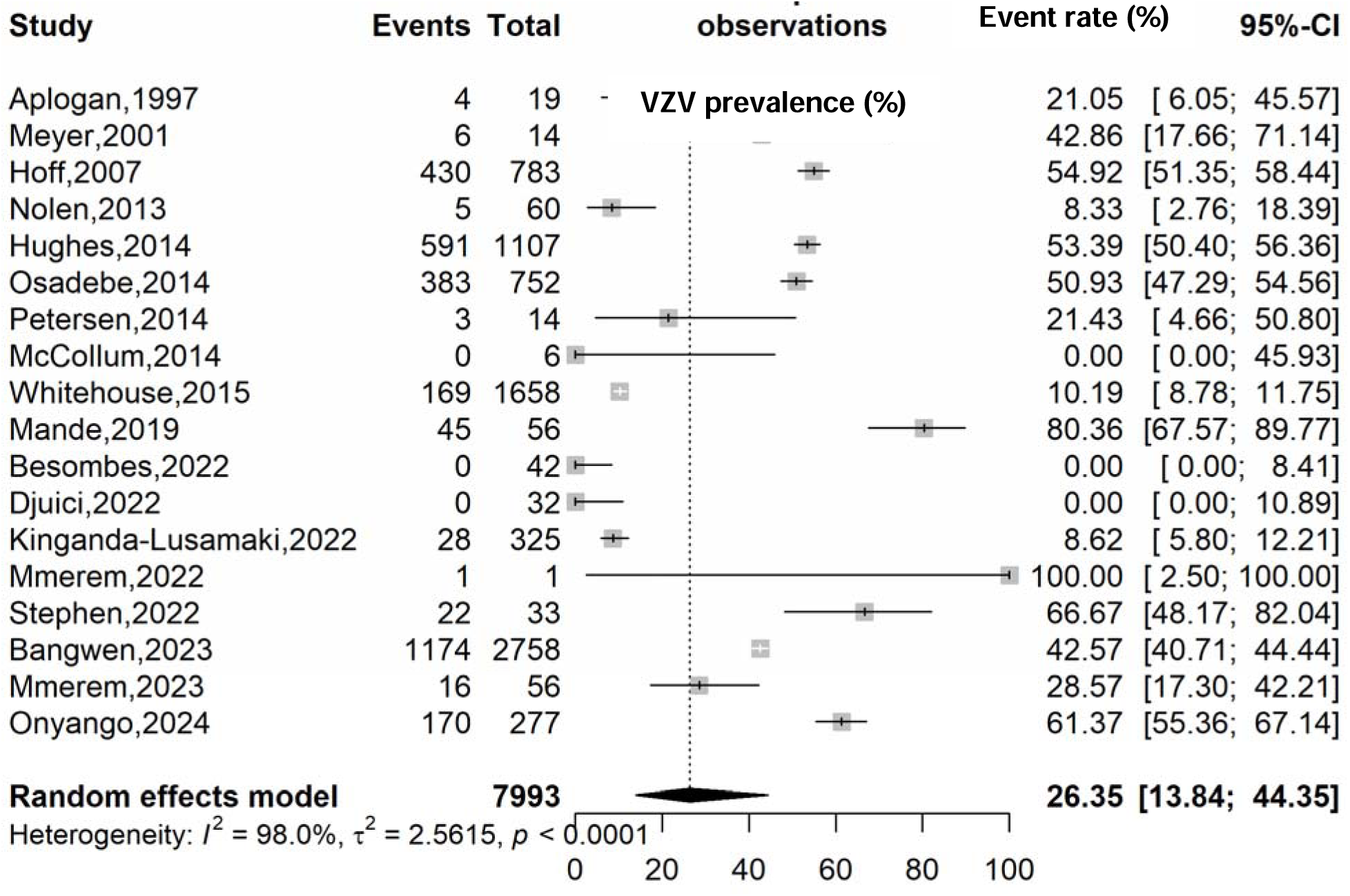
Prevalence of varicella-zoster virus (VZV) infections in Africa

Subgroup analyses indicated higher pooled VZV prevalence in studies conducted before 2022 (31.50%; 95% CI: 16.81–51.12; 4,469 participants; n = 10 studies) compared to those published from 2022 onward (19.83%; 95% CI: 4.14–58.59; 3,524 participants; n = 8 studies) although this difference was not statistically significant (*p* = 0.529). In contrast, notable differences were observed by countries (*p* = 0.019) and WHO region (*p* = 0.002). The highest prevalence was reported in Kenya (61.37%; 95% CI: 55.36–67.14; 277 participants; n = 1 study) and Nigeria (50.37%; 95% CI: 24.03–76.51; 90 participants; n = 3 studies), while lower prevalence estimates were seen in the Democratic Republic of Congo (29.38%; 95% CI: 16.64–46.43; 7,552 participants; n = 12 studies) and across Central Africa overall (19.64%; 95% CI: 8.53–39.06; 7,626 participants; n = 14 studies) (Table 4).

**Table 4.** Subgroup meta-analysis of the pooled prevalence of varicella-zoster virus infections in Africa.

| Subgroup | Sample size | Prevalence<sup>1</sup> (%) | 95% CI limits<sup>1</sup> |  | Number of reports | Heterogeneity statistic<sup>1</sup> |  | Subgroup difference p-value |
| --- | --- | --- | --- | --- | --- | --- | --- | --- |
|  |  |  | Lower | Upper |  | I<sup>2</sup>(%) | p-value |  |
| **Study period (year)** |  |  |  |  |  |  |  | 0.529 |
| Before 2022 | 4,469 | 31.50 | 16.81 | 51.12 | 10 | 98.7 | □ 0.001 |  |
| 2022 + | 3,524 | 19.83 | 4.14 | 58.59 | 8 | 95.6 | □ 0.001 |  |
| **Study design** |  |  |  |  |  |  |  | 0.562 |
| Cross-sectional | 7,930 | 26.67 | 13.29 | 46.32 | 15 | 98.4 | □ 0.001 |  |
| Cohort | 56 | 28.57 | 17.30 | 42.21 | 1 | - | - |  |
| Case report | 7 | 0.00 | 0.00 | 99.99 | 2 | 0.0 | 0.999 |  |
| **Study setting** |  |  |  |  |  |  |  | 0.230 |
| Community | 5,113 | 24.99 | 11.98 | 44.91 | 13 | 98.6 | □ 0.001 |  |
| Hospital | 90 | 50.37 | 24.03 | 76.51 | 3 | 82.7 | 0.003 |  |
| Community and hospital | 2,790 | 4.28 | 0.02 | 91.54 | 2 | 0.0 | 0.999 |  |
| **Participant** |  |  |  |  |  |  |  |  |
| General population | 7,919 | 28.49 | 14.06 | 49.24 | 16 | 98.2 | □ 0.001 |  |
| Healthcare worker | 14 | 21.43 | 4.66 | 50.80 | 1 | - | - |  |
| General population and healthcare worker | 60 | 8.33 | 2.76 | 18.39 | 1 | - | - |  |
| **Country** |  |  |  |  |  |  |  | 0.019 |
| DRC | 7,552 | 29.38 | 16.64 | 46.43 | 12 | 98.6 | □ 0.001 |  |
| Nigeria | 90 | 50.37 | 24.03 | 76.51 | 3 | 82.7 | 0.003 |  |
| CAR | 42 | 0.00 | 0.00 | 8.41 | 1 | - | - |  |
| Cameroon | 32 | 0.00 | 0.00 | 10.89 | 1 | - | - |  |
| Kenya | 277 | 61.37 | 55.36 | 67.14 | 1 | - | - |  |
| **WHO zone<sup>2</sup>** |  |  |  |  |  |  |  | 0.002 |
| Western | 90 | 50.37 | 24.03 | 76.51 | 3 | 82.7 | 0.003 |  |
| Eastern | 277 | 61.37 | 55.36 | 67.14 | 1 | - | - |  |
| Central | 7,626 | 19.64 | 8.53 | 39.06 | 14 | 98.4 | □ 0.001 |  |

<sup>1</sup> Random effects model; CI: Confidence interval; WHO: World Health Organization; <sup>2</sup> Western: Nigeria; Eastern: Kenya; Central: Cameroon, CAR = Central African Republic, and DRC = Democratic Republic of Congo

### 3.6. Meta-regression analysis

Meta-regression analysis showed that for VZV–mpox coinfection, studies from the Western WHO zone remained significantly associated with higher prevalence (β = 4.42; *p* = 0.001). For all other study outcomes, no characteristic was identified as significant source of heterogeneity (Table 5).

**Table 5.** Meta-regression analysis of pooled estimates of prevalence of VZV, HIV infections among confirmed mpox cases in Africa.

| Estimate | Moderator | Univariate |  | Multivariate |  |
| --- | --- | --- | --- | --- | --- |
|  |  | Unadjusted coefficient (β) | p-value | Adjusted coefficient (β) | p-value |
| VZV-mpox coinfection |  |  |  |  |  |
| Study year | Year | 0.0174 | 0.781 | -0.0950 | 0.194 |
| Design | Cross-sectional vs. Others | -1.1099 | 0.311 | 0.6529 | 0.594 |
| Participant | Healthcare worker vs. General population | 0.0068 | 0.997 | 1.3995 | 0.401 |
| Setting<sup>1</sup> | Hospital vs. Others | 2.1685 | 0.005 | - | - |
| WHO zone<sup>2</sup> | Central | Ref | - | Ref | - |
|  | Western | 0.4376 | 0.724 | 4.4212 | **0.001** |
|  | Eastern | 2.3678 | 0.007 | 2.2230 | 0.149 |
| Disease burden | Number of confirmed cases | 0.0001 | 0.950 | 0.0016 | 0.186 |
| HIV-mpox coinfection |  |  |  |  |  |
| Study year | Year | -0.0130 | 0.878 | -0.0942 | 0.467 |
| Design | Cross-sectional vs. Others | -1.6132 | 0.155 | 0.5173 | 0.549 |
| Setting | Hospital vs. Others | 2.0428 | 0.025 | 0.2895 | 0.813 |
| WHO zone<sup>2</sup> | Central | Ref | - | Ref | - |
|  | Western | 2.8120 | < 0.001 | 1.5620 | 0.219 |
|  | Eastern | 1.0304 | 0.348 | 0.8276 | 0.509 |
| Disease burden | Number of confirmed cases | -0.0040 | < 0.001 | -0.0024 | 0.051 |
| VZV infection |  |  |  |  |  |
| Study year | Year | 0.0002 | 0.997 | -0.0527 | 0.417 |
| Design | Cross-sectional vs. Others | 0.0782 | 0.941 | 1.2438 | 0.401 |
| Participant | General population vs. Others | 1.2180 | 0.239 | 0.5978 | 0.645 |
| Setting | Community vs. Others | -0.3576 | 0.655 | 1.1903 | 0.471 |
| WHO zone<sup>2</sup> | Central | Ref | - | Ref | - |
|  | Western | 1.1848 | 0.217 | 3.6982 | 0.076 |
|  | Eastern | 1.5571 | 0.261 | 2.1144 | 2.1144 |
| Disease burden | Number of confirmed cases | 0.0002 | 0.717 | 0.0007 | 0.268 |

<sup>1</sup> Redundant predictor dropped from the model; <sup>2</sup> Western: Nigeria; Eastern: Kenya; Central: Cameroon, Central African Republic, and Democratic Republic of Congo; WHO: World Health Organization

### 3.7. Clinical pattern of confirmed mpox cases by genotypic category in Africa

Clinical manifestations of mpox varied significantly by viral clade. Rash was nearly universal across clades (>96%) and did not differ by genotype (*p* = 0.118). In contrast, systemic symptoms (fever, lymphadenopathy, sore throat, myalgia, fatigue, headache, and chills) were significantly more frequent in clades I and Ia than in clade II (all *p* < 0.01). Fever and lymphadenopathy were almost universal in clades I/Ia (≥90–100%) but occurred less frequently in clade II, particularly fever (16.8%).

Mucocutaneous manifestations also differed by clade, with oral lesions, conjunctivitis, palm and sole lesions, and genital lesions occurring more often in clades I and Ia (*p* < 0.001 for most). Gastrointestinal and respiratory symptoms showed no significant clade-specific differences (*p* > 0.05). Overall, clades I and Ia were associated with more pronounced systemic disease, whereas clade II showed a milder systemic profile. Similarly, Congo basin clades case significantly revealed a richer clinical pattern compared to West Africa clade (Table 6 & 7).

**Table 6.**
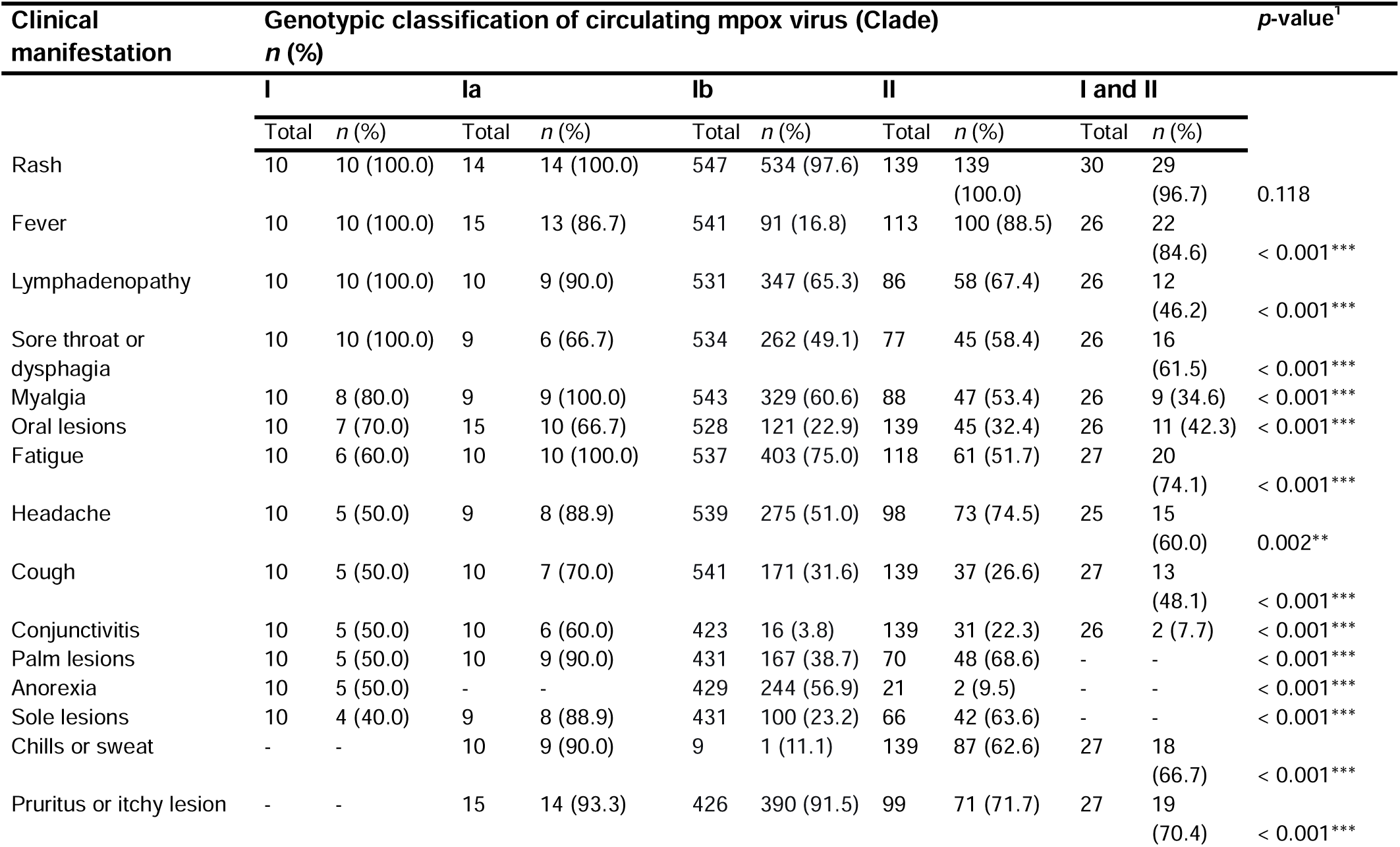
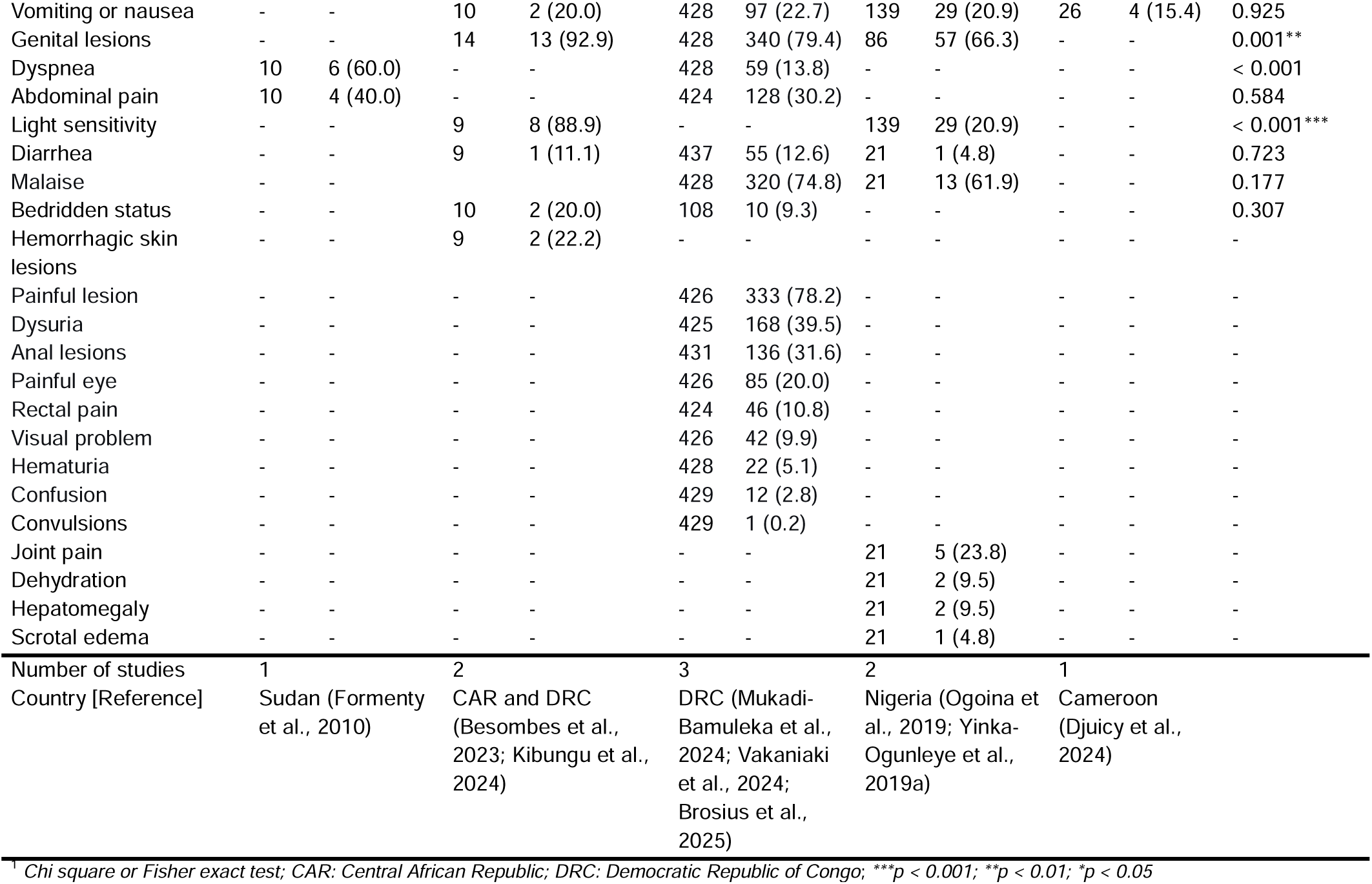
Clinical pattern of confirmed mpox cases by genotypic category in Africa.

**Table 7.**
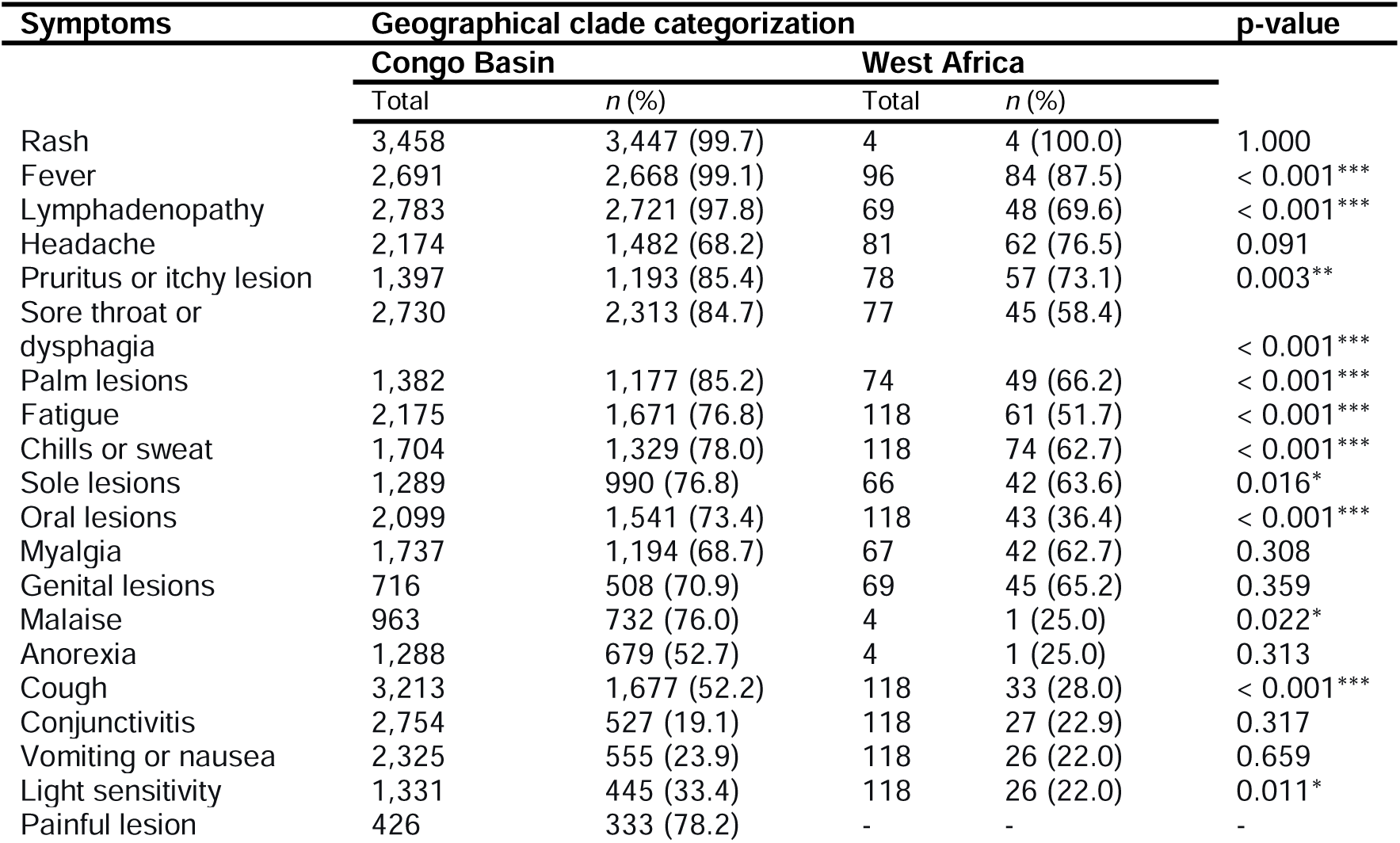
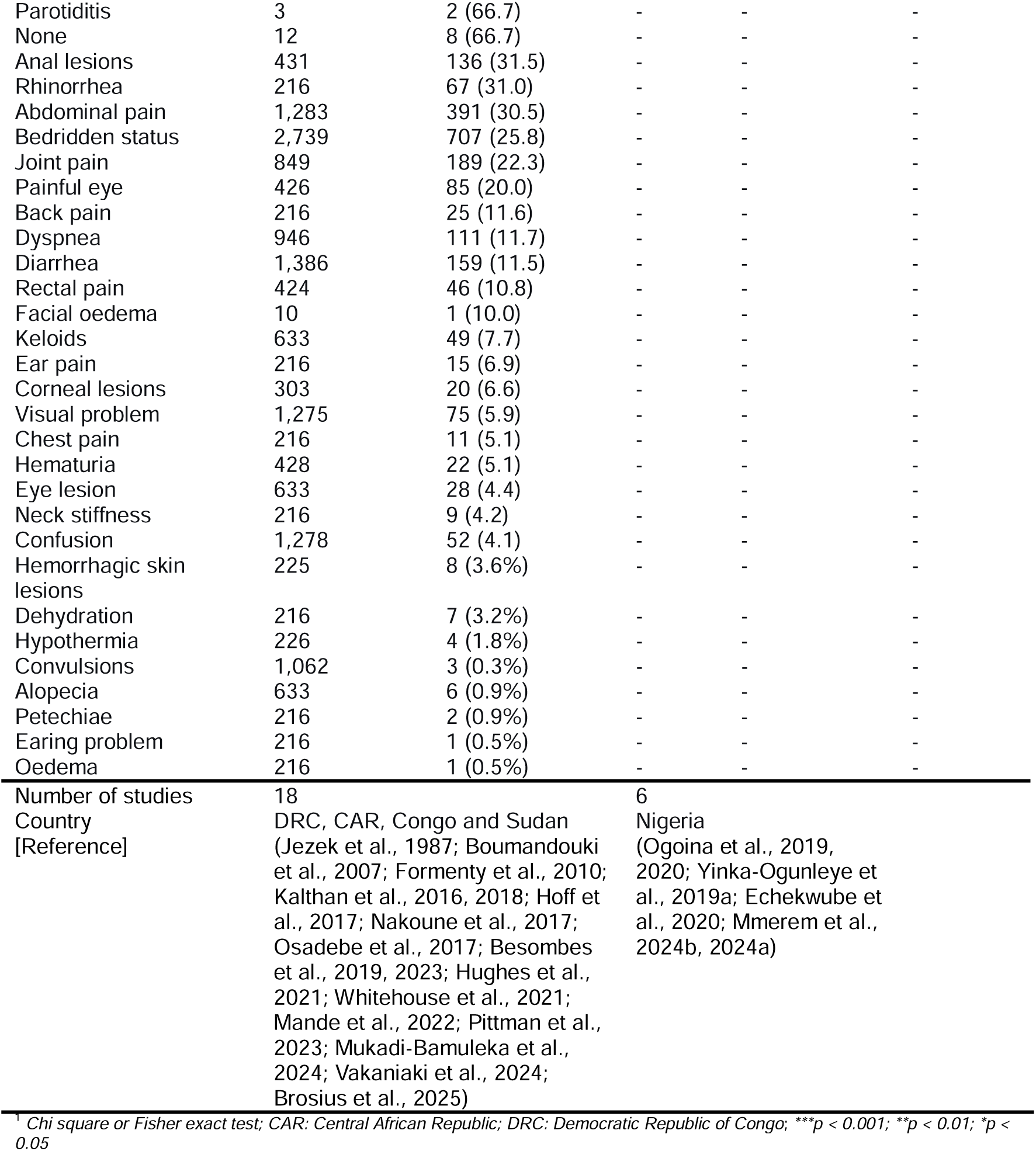
Clinical profile of confirmed mpox cases by geographical clade classification in Africa.

## 4. Discussion

This systematic review and meta-analysis aimed to determine the coinfection rate of Mpox and VZV, as well as the clinical profile of Mpox cases in Africa. It compiled data from 27 studies across Africa.

### 4.1. VZV-mpox coinfection

Our meta-analysis of 2,691 confirmed mpox cases in Africa showed a pooled VZV coinfection rate of 10.23% (95% CI: 2.95–29.93). This indicates that VZV coinfection is a common feature of mpox epidemiology, especially in endemic areas and during outbreaks (Hoff et al., 2017; Stephen et al., 2022, 2022). Overall, these findings show significant variability in estimates across different studies (*I*^2^ = 67%) and the estimate should be interpreted with caution, although this could be explained by different study settings, study designs, and study populations.

The subgroup analysis revealed a significantly higher prevalence (*p* < 0.001) in hospital-based studies (60.66%) compared to community-based studies (7.60%). This difference may be attributed to the fact that patients admitted for mpox and coinfected with VZV might present with more pronounced disease severity or atypical clinical presentations, which lead the patient to seek hospital care (Benites-Zapata et al., 2022). Furthermore, the clinical similarities between VZV and mpox skin lesions may lead to misclassification, potentially affecting prevalence estimates in a hospital setting (Yinka-Ogunleye et al., 2019b; Bourner et al., 2024).

Regarding the regional distribution of VZV–mpox coinfection, the highest prevalences were reported in West Africa (60.66%), followed by East Africa (15.38%), while Central Africa showed the lowest rates (4.28%). The main mpox hotspots in these regions are Nigeria, Uganda, and the DRC, respectively, which together account for more than 90% of mpox-related deaths in Africa (Cheuyem et al., 2025b; An Update of Monkeypox Outbreak in Nigeria, n.d.; Multi-country monkeypox outbreak: situation update, n.d.). This regional variation may be due to differences in surveillance systems and diagnostic capacities. Nigeria and Uganda have improved their surveillance and diagnostic capabilities, which help identify and report cases (Boboye Onduku, 2019; Reinforcing outbreak response systems to curb mpox in Uganda | WHO | Regional Office for Africa, 2025). In contrast, despite being an endemic focus, the DRC faces significant surveillance limitations, especially in remote regions, leading to underreporting of coinfected cases (Adigun et al., 2024). Furthermore, differences in access to diagnostic tools, especially PCR testing, directly affect the reported prevalence estimates across regions (Bourner et al., 2024).

### 4.2. HIV-mpox coinfection

The overall prevalence of HIV coinfection among patients with mpox in our analysis was 6.55% (95% CI: 2.36–16.90). Indeed, several studies have confirmed the presence of HIV–mpox coinfection, especially in Nigeria and the DRC (Echekwube et al., 2020; Mmerem et al., 2024b; Cheuyem et al., 2025a). This implies that HIV–mpox coinfection does occur and may reach variable proportions depending on the context. The high heterogeneity observed suggests that we should interpret the pooled estimate with caution.

The WHO regional analysis mirrors the country-level findings, with the highest prevalence significantly observed (*p* < 0.001) in the West Africa region (21.19%), followed by East Africa (4.55%) and Central Africa (1.21%). In contrast, a recent meta-analysis examining the geographic and temporal variation of mpox patients living with HIV worldwide reported substantially higher HIV–mpox coinfection prevalences among mpox cases in Europe (41%) and North America (52%) (Gandhi et al., 2023). These findings highlight significant geographic differences in HIV–mpox coinfection patterns and suggest that screening and management strategies should be customized to the local epidemiological context.

At the country level, a higher prevalence was observed in Nigeria (21%), an intermediate prevalence in Burundi (4.6%), and a lower prevalence in the DRC (1.2%). This variation may be explained by several factors, including the decreasing gradient of HIV prevalence across these countries (Nigeria > Burundi > DRC), which mechanically influences the risk of coinfection (Prevalence of HIV, total (% of population ages 15-49), n.d.). Additionally, mpox epidemiological profiles vary greatly across different settings. In the DRC, mpox transmission has mainly been driven by zoonotic and intrafamilial contacts, mostly impacting rural populations and children, among whom HIV rates are low. Conversely, recent outbreaks in Nigeria have involved ongoing human-to-human transmission, including sexual contact, which overlaps with HIV transmission networks (McCollum et al., 2015; Ogoina et al., 2020).

A significant difference was also observed according to study setting (*p* = 0.007), with hospital-based studies reporting a substantially higher prevalence of HIV–mpox co-infection (15.97%) compared with community-based studies (1.75%). These findings are consistent with a recent meta-analysis showing that individuals co-infected with HIV and mpox had a significantly higher likelihood of hospitalization than those infected with mpox alone (OR = 1.85) (Taha et al., 2024). Similarly, another meta-analysis reported a pooled 56.6% increased risk of hospitalization among HIV-positive mpox cases compared with HIV-negative individuals (95% CI: 18.0–107.7%) (Shabil et al., 2025). These findings support the biological plausibility that people living with HIV, due to immune system impairment, are more susceptible to developing severe or complicated forms of mpox, which increases the likelihood of hospital-based case detection (Echekwube et al., 2020; Mmerem et al., 2024b).

The study design significantly influenced the estimates of HIV–mpox coinfection. Case reports reported the highest coinfection rate (100.0%), followed by case series (75.0%), whereas cross-sectional and cohort studies reported much lower rates (ranging from 3.99% to 4.85%). The markedly higher HIV–mpox coinfection rates observed in case reports and case series are likely attributable to the focus of these studies on severe, atypical, or hospitalized cases, in which comorbidities, particularly HIV infection, are more frequently reported (McCollum et al., 2015; Echekwube et al., 2020; Mmerem et al., 2024b).

### 4.3. Mpox clinical presentation

In this study, skin rash was the predominant clinical manifestation, observed in more than 96% of cases, which is consistent with the World Health Organization (WHO) case definitions that consider rash to be the cardinal symptom of suspected mpox (WHO, 2024). This finding confirms the robustness of current clinical criteria for case identification, particularly in resource-limited settings.

Beyond cutaneous manifestations, we observed a high frequency of systemic symptoms, including fever, lymphadenopathy, sore throat, myalgia, fatigue, headache, and chills, with a significantly higher prevalence in clades I and Ia compared with clade II. This distribution is consistent with historical data suggesting a more severe clinical presentation associated with clade I, characterized by marked systemic involvement. Indeed, large clinical series and outbreak investigations conducted in Africa have reported a high prevalence of generalized symptoms among patients infected with this clade, supporting the hypothesis of intrinsically higher virulence compared with clade II (Hutin et al., 2001; Besombes et al., 2023; Djuicy et al., 2024).

Our supporting evidence is strengthened by enhanced surveillance studies and analyses of previous outbreaks, which show that infections attributed to clades I and Ia are often accompanied by fever, chills, severe fatigue, headache, and lymphadenopathy (Formenty et al., 2010; Stephen et al., 2022). However, the predominance of these non-specific symptoms complicates the differential diagnosis, particularly in endemic regions where febrile illnesses such as malaria, varicella-zoster virus (VZV) infection, and other viral or bacterial infections are highly prevalent (Viral Febrile Illnesses and Emerging Pathogens, 2020; Latest information on diseases from Africa CDC, n.d.). This clinical overlap might lead to delays in diagnosis or missed mpox cases, especially when the rash is absent or appears late.

Despite the overall high quality of the included research, significant heterogeneity remained across several analyses, highlighting significant gaps in surveillance coverage, diagnostic capabilities, and study design throughout the continent. In addition, some estimates were based on few studies with wide confidence intervals; therefore, these subgroup findings should be interpreted cautiously.

In Africa, VZV and HIV are found to be coinfect several mpox patient which can result to more severe disease and uncommon and pronounced clinical presentations. The fact that coinfections are significantly more common in hospital settings suggests that they may play a role in the severity forms of mpox that need specialized treatment. The clinical signs varied considerably depending on the viral clade, with clades I and Ia exhibiting more systemic involvement than clade II. These findings support enduring worries about the increased intrinsic pathogenicity of Congo Basin clades and emphasize the additional benefit of combining clinical and genetic data in mpox surveillance and response plans. Improving risk classification, clinical treatment, and outbreak readiness requires strengthening integrated monitoring systems, increasing routine HIV and VZV screening among mpox patients, and improving genomic characterization of circulating strains. To lessen mpox-related morbidity and promote efficient and equitable control strategies throughout Africa, it will be necessary to produce strong, context-specific primary evidence.

## Supporting information

Addition Files 1

## Data availability statement

The sources of data supporting this systematic review are available in the reference. All data generated or analyzed during this study are included in this published article and supplemental material.

## Author contribution

Conceptualization: FZLC; Methodology: FZLC, CA, LBKB, and AEM; Data curation: FZLC, CA, AEM; Software: FZLC; Validation: F.Z.L.C; Formal analysis: F.Z.L.C; Funding acquisition: None; Investigation: FZLC, CA, LBKB, and AEM; Resources: All authors; Writing—original draft preparation: FZLC, CA, LBKB, RT and CNM; Writing—review and editing: All authors; Visualization: FZLC: Supervision: FZLC; Project administration: FZLC; All authors have read and agreed to the published version of the manuscript.

## Funding source

This research did not receive any specific grant from funding agencies in the public, commercial or not-for-profit sectors.

## Acknowledgements

None.

## Conflict of interest

The authors declare that the research was conducted in the absence of any commercial or financial relationships that could be construed as a potential conflict of interest.

## Abbreviations

CAR: Central African Republic
*CI*: Confidence interval
*DRC*: Democratic Republic of Congo
GP: General population
*HIV*: Human immunodeficiency virus/acquired immunodeficiency syndrome
*MeSH*: Medical subject headings
*PCR*: Polymerase chain reaction
*PRISMA*: Preferred reporting items for systematic reviews and meta-analysis
VZV: Varicella-zoster virus.

## Notes

### Competing Interest Statement

The authors have declared no competing interest.

### Funding Statement

This study did not receive any funding

### Author Declarations

The sources of data supporting this systematic review are available in the reference.

