## Additional Files 2 for "Mpox coinfections and clinical manifestation in Africa: a systematic review and meta-analysis"

**Study period**

**HIV coinfection rate (%)**

**Event rate (%)**

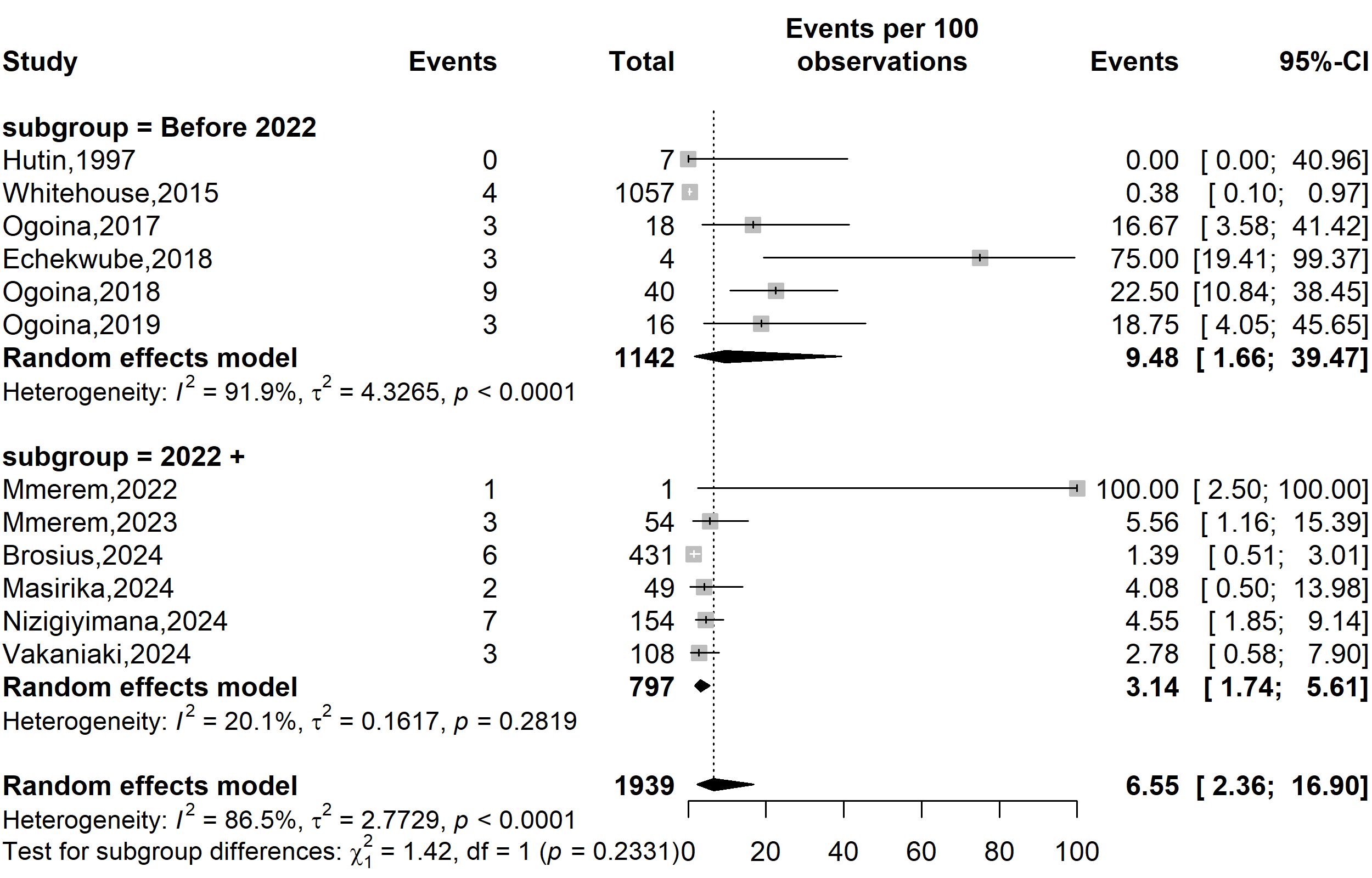

**Supplementary Fig. 1** Prevalence of human immunodeficiency virus (HIV) coinfections among confirmed mpox cases in Africa by study period

**Country**

**Event rate (%)**

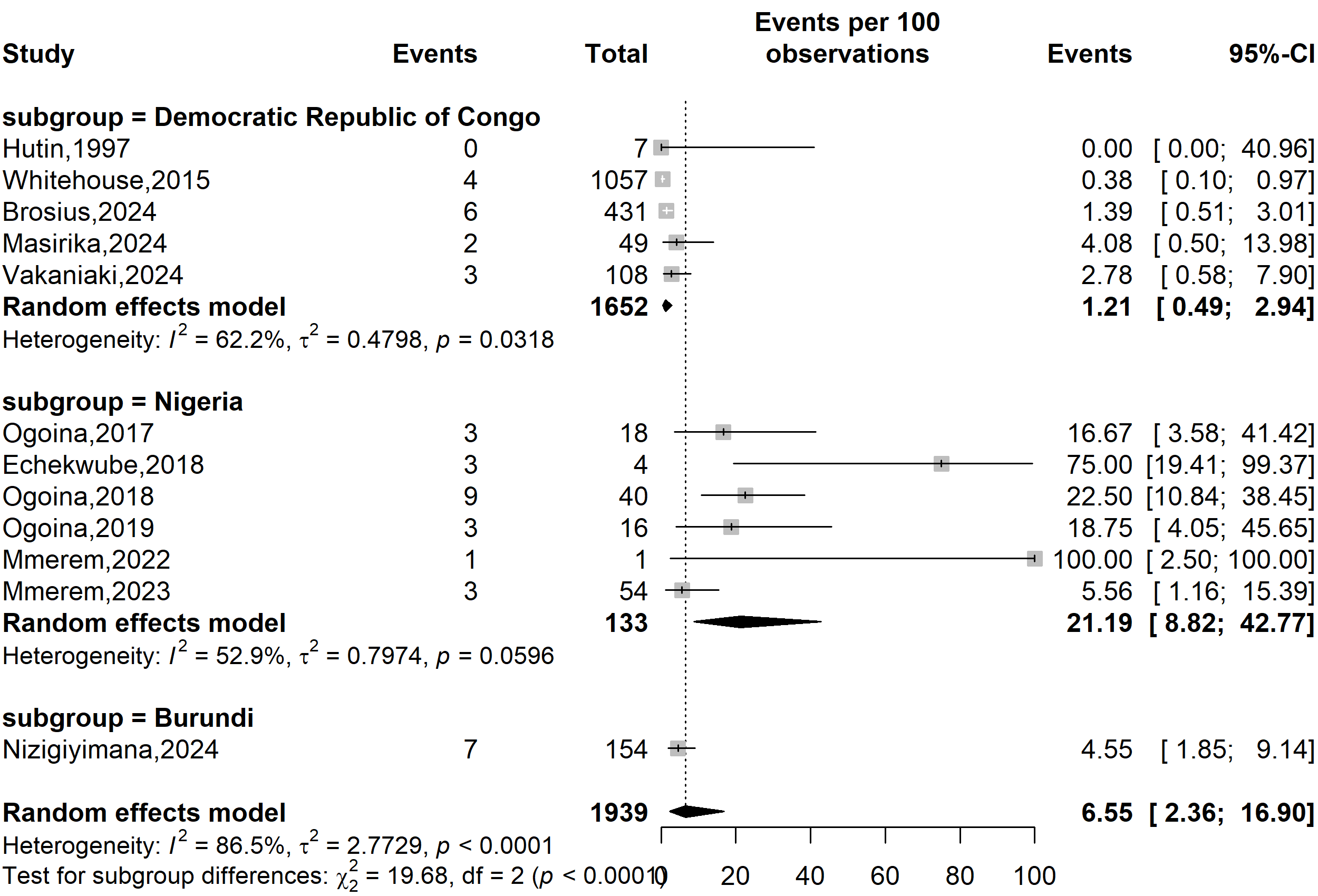

**HIV coinfection rate (%)**

**Supplementary Fig. 2** Prevalence human immunodeficiency virus (HIV) coinfections among confirmed mpox cases in Africa by country

**Study design**

**HIV coinfection rate (%)**

**Event rate (%)**

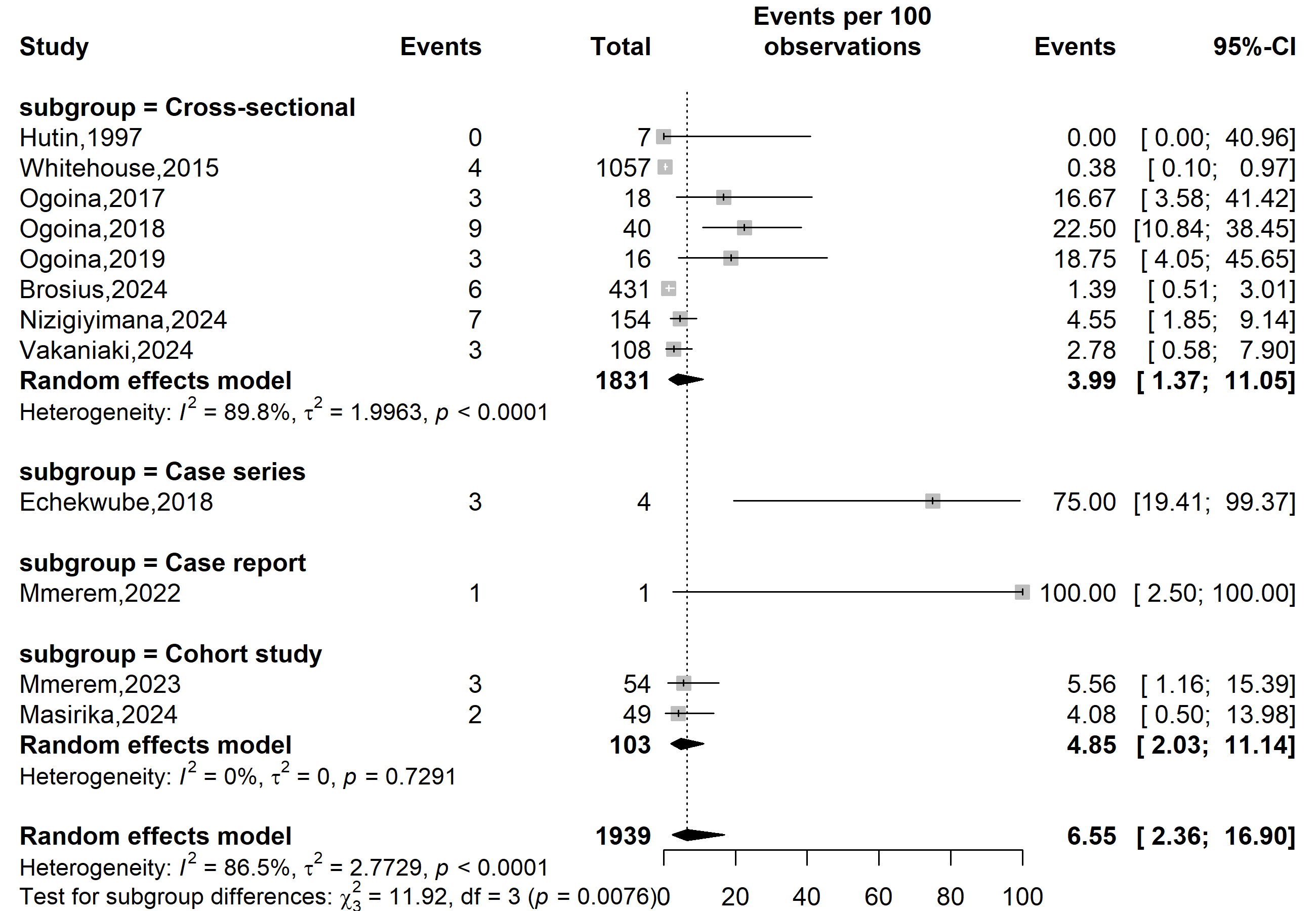

**Supplementary Fig. 3** Prevalence human immunodeficiency virus (HIV) coinfections among confirmed mpox cases in Africa by study design

**Study design**

**Event rate (%)**

**HIV coinfection rate (%)**

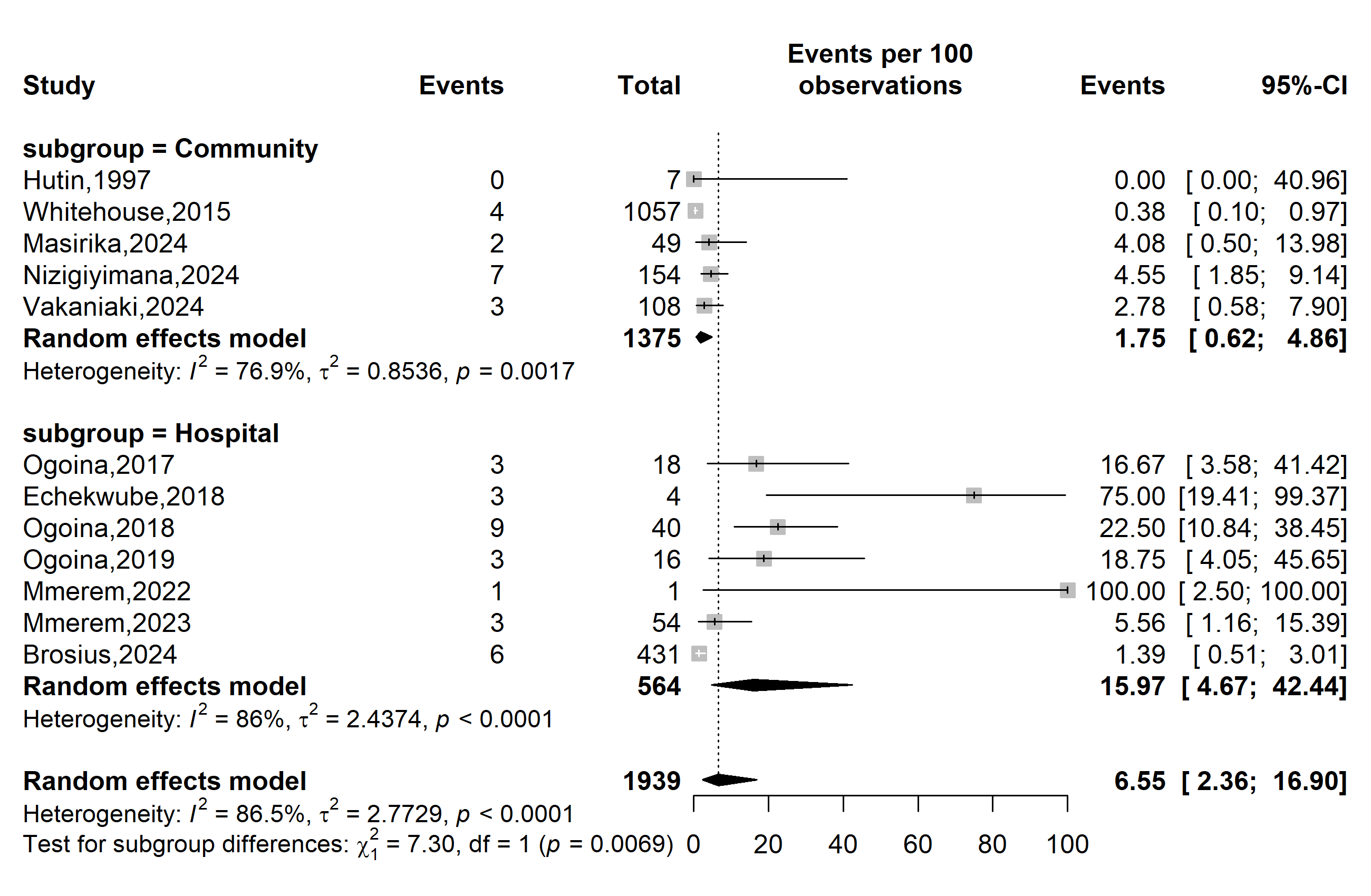

**Supplementary Fig. 4** Prevalence human immunodeficiency virus (HIV) coinfections among confirmed mpox cases in Africa by study setting

**Study WHO Afro region**

**Event rate (%)**

**HIV coinfection rate (%)**

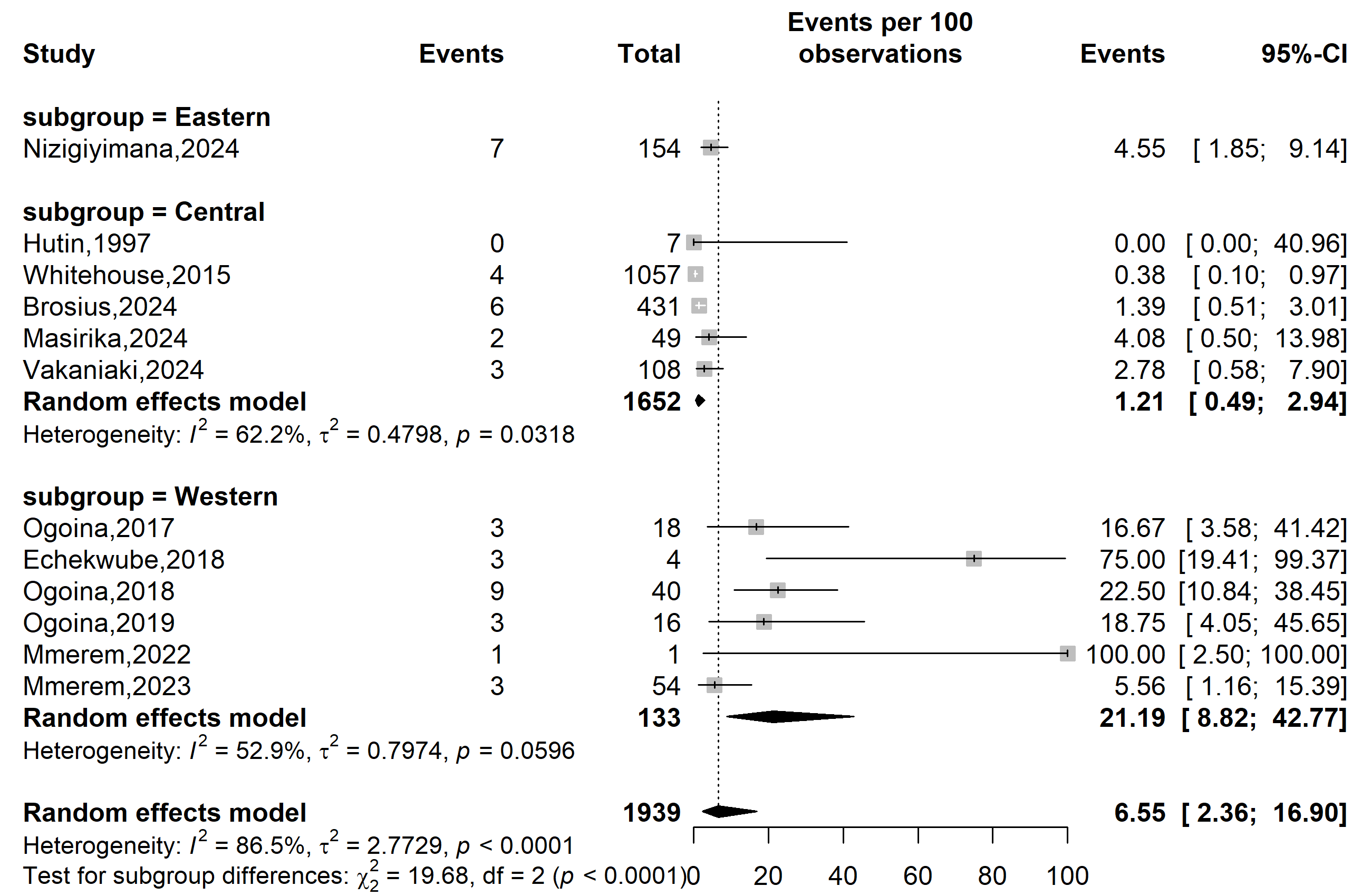

**Supplementary Fig. 5** Prevalence human immunodeficiency virus (HIV) coinfections among confirmed mpox cases in Africa by WHO Afro region

**Publication bias**

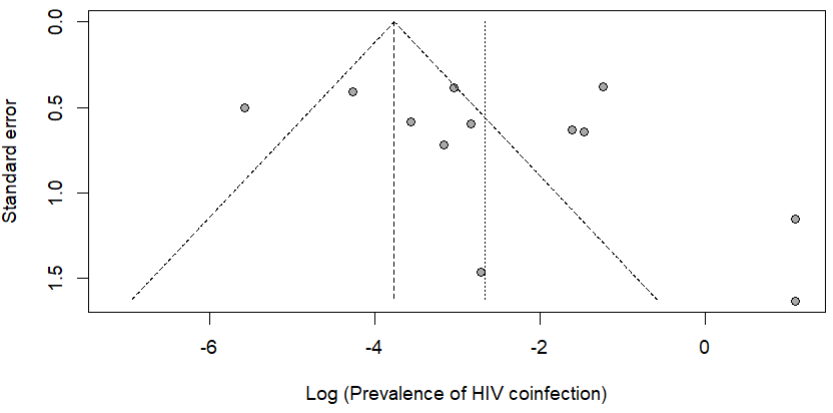

Egger’s test *p*-value = 0.322

Begg’s test *p*-value = 0.131

**Supplementary Fig. 6** Funnel plot displaying the pseudo 95% confidence limits and tests assessing the publication bias of studies included

**Sensitivity analysis**

**Event rate (%)**

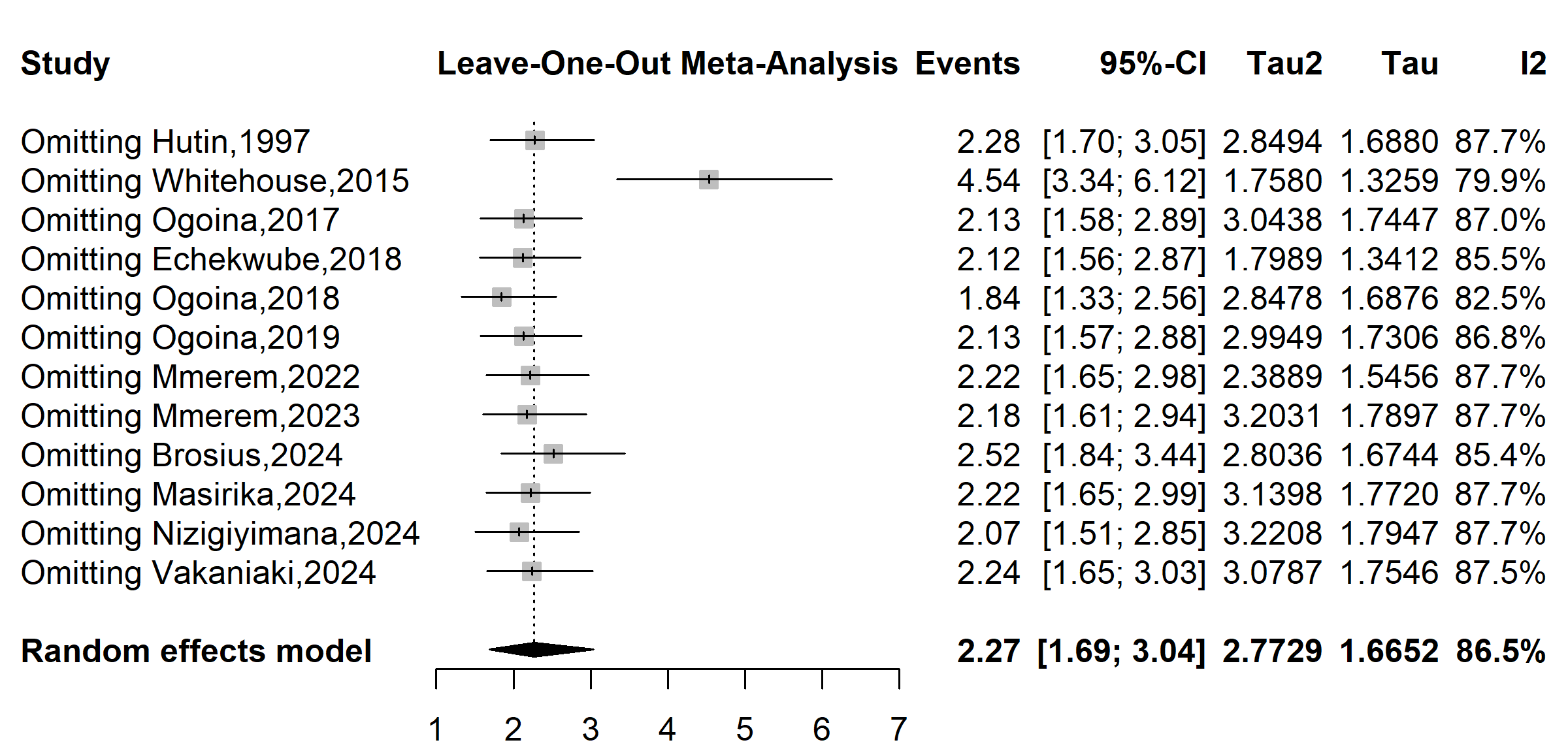

**Supplementary Fig. 7** Sensitivity analysis of the prevalence human immunodeficiency virus (HIV) coinfections among confirmed mpox cases in Africa
