## Additional Files 3 for "Mpox coinfections and clinical manifestation in Africa: a systematic review and meta-analysis"

**Study period**

**VZV prevalence (%)**

**Event rate (%)**

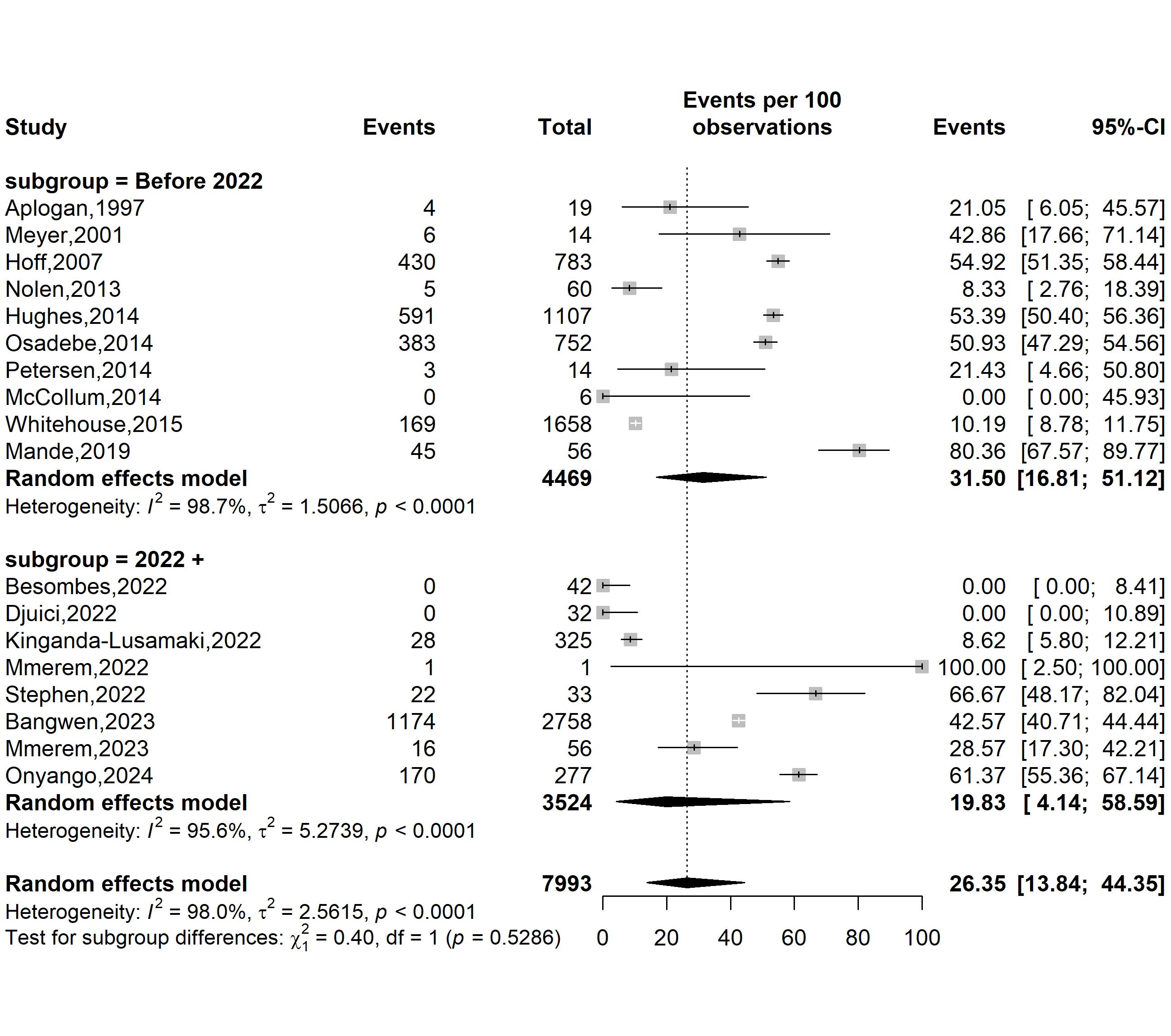

**Supplementary Fig. 1** Prevalence varicella-zoster virus (VZV) infections in Africa by study period

**Country**

**Event rate (%)**

**VZV prevalence (%)**

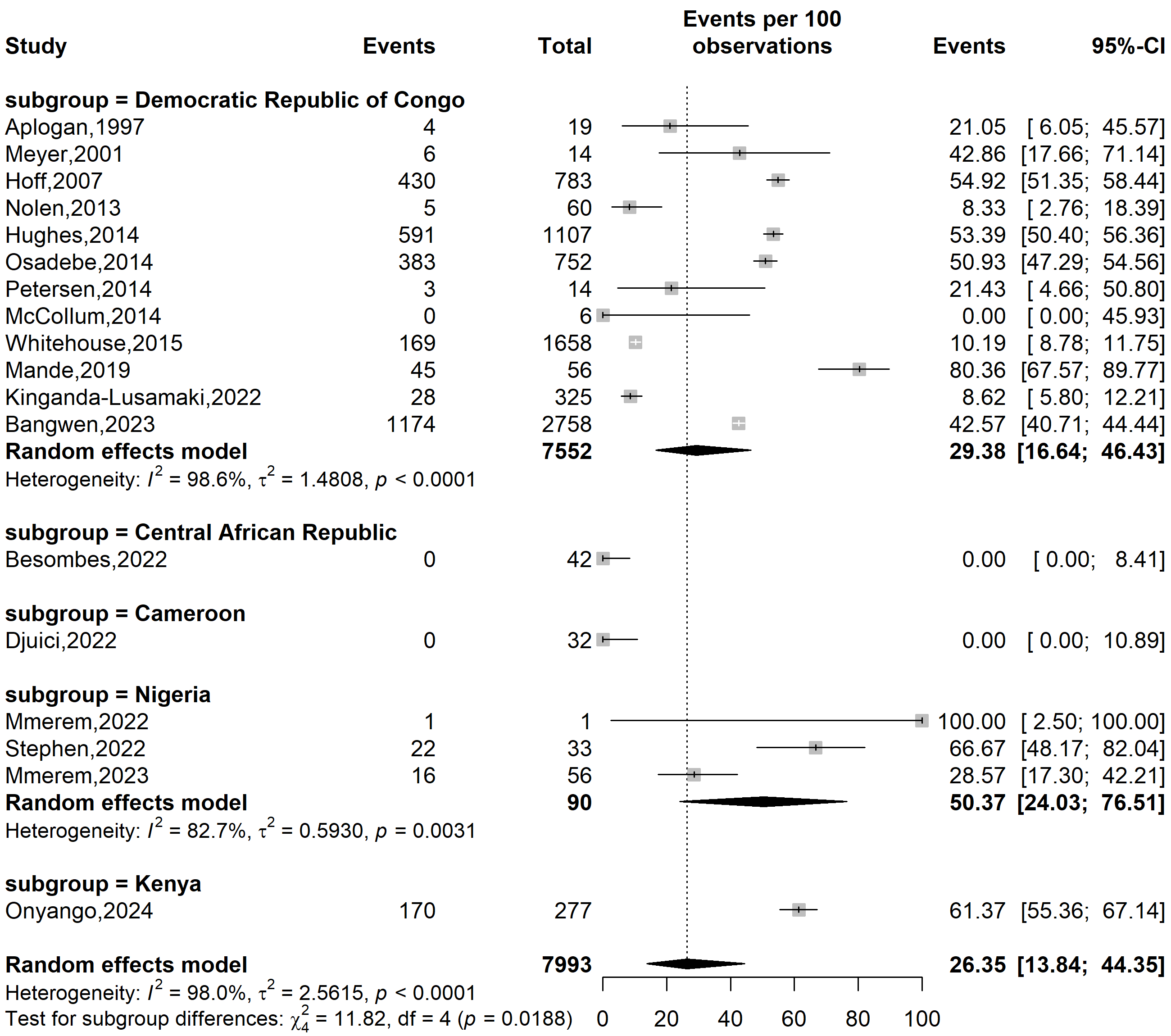

**Supplementary Fig. 2** Prevalence varicella-zoster virus (VZV) infections in Africa by country

**Study design**

**VZV prevalence (%)**

**Event rate (%)**

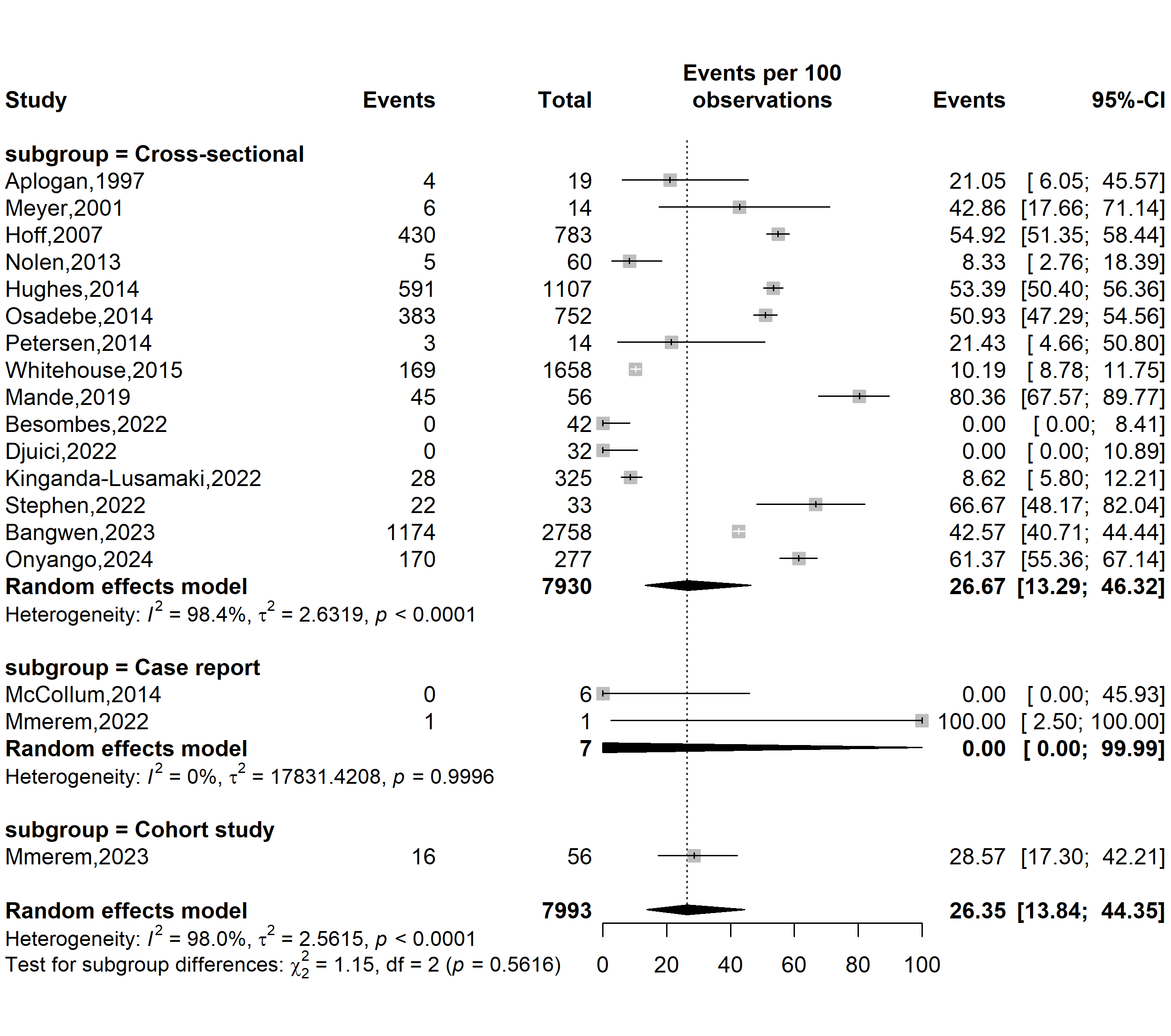

**Supplementary Fig. 3** Prevalence varicella-zoster virus (VZV) infections in Africa by study design

**Study setting**

**VZV prevalence (%)**

**Event rate (%)**

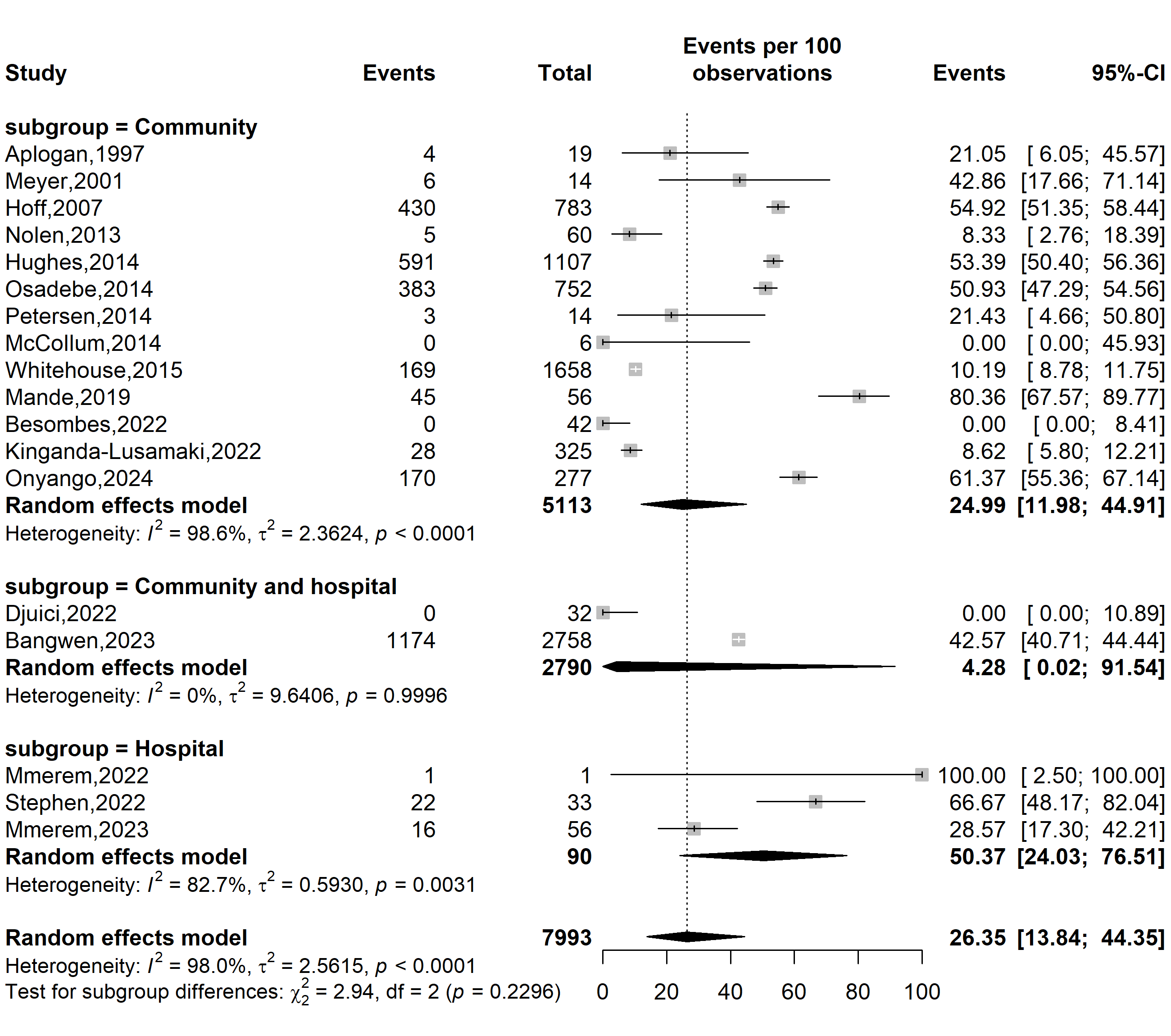

**Supplementary Fig. 4** Prevalence varicella-zoster virus (VZV) infections in Africa by study setting

**Study participant**

**Event rate (%)**

**VZV prevalence (%)**

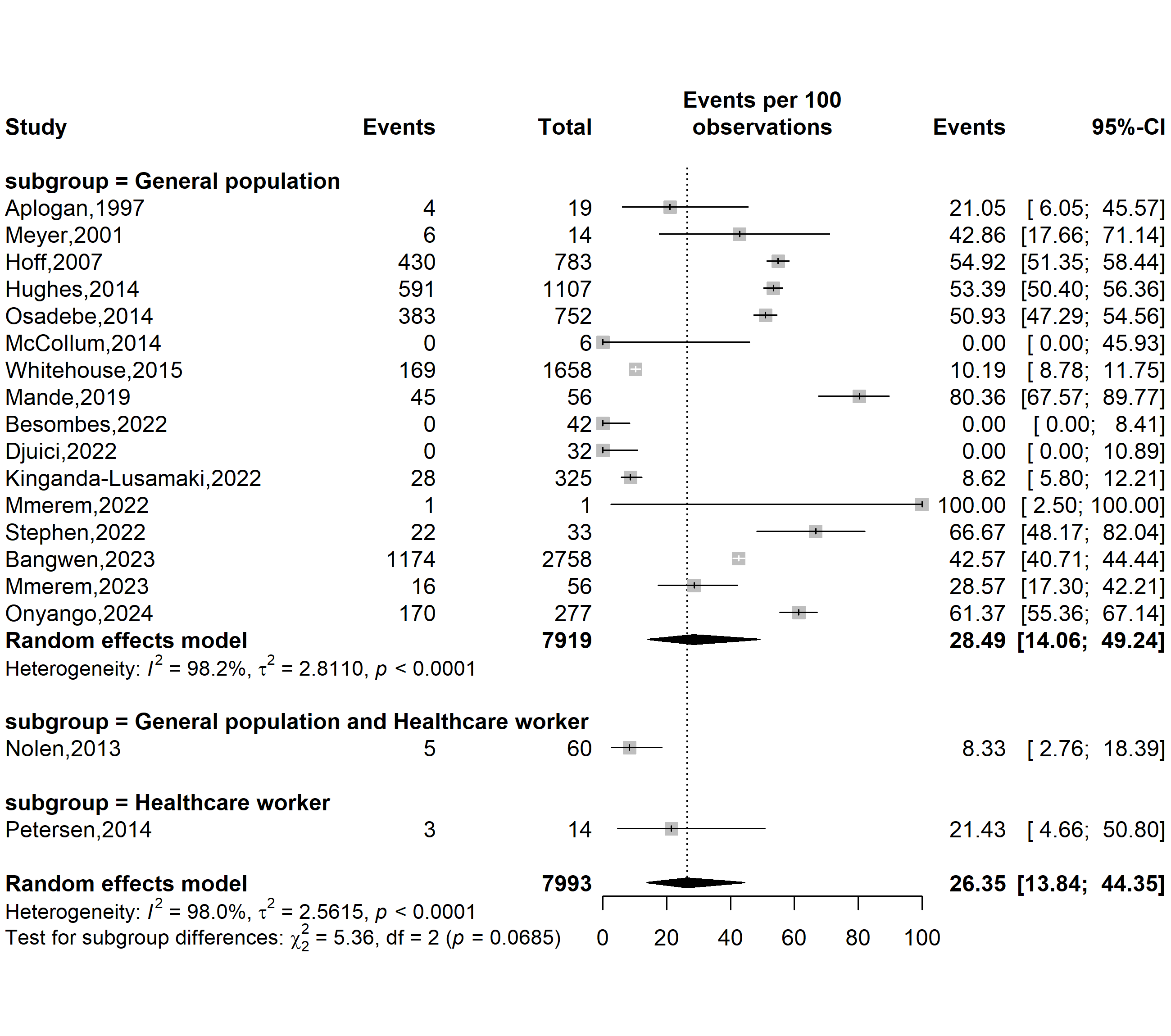

**Supplementary Fig. 5** Prevalence varicella-zoster virus (VZV) coinfections among confirmed mpox cases in Africa by study participants

**Study WHO Afro region**

**VZV prevalence (%)**

**Event rate (%)**

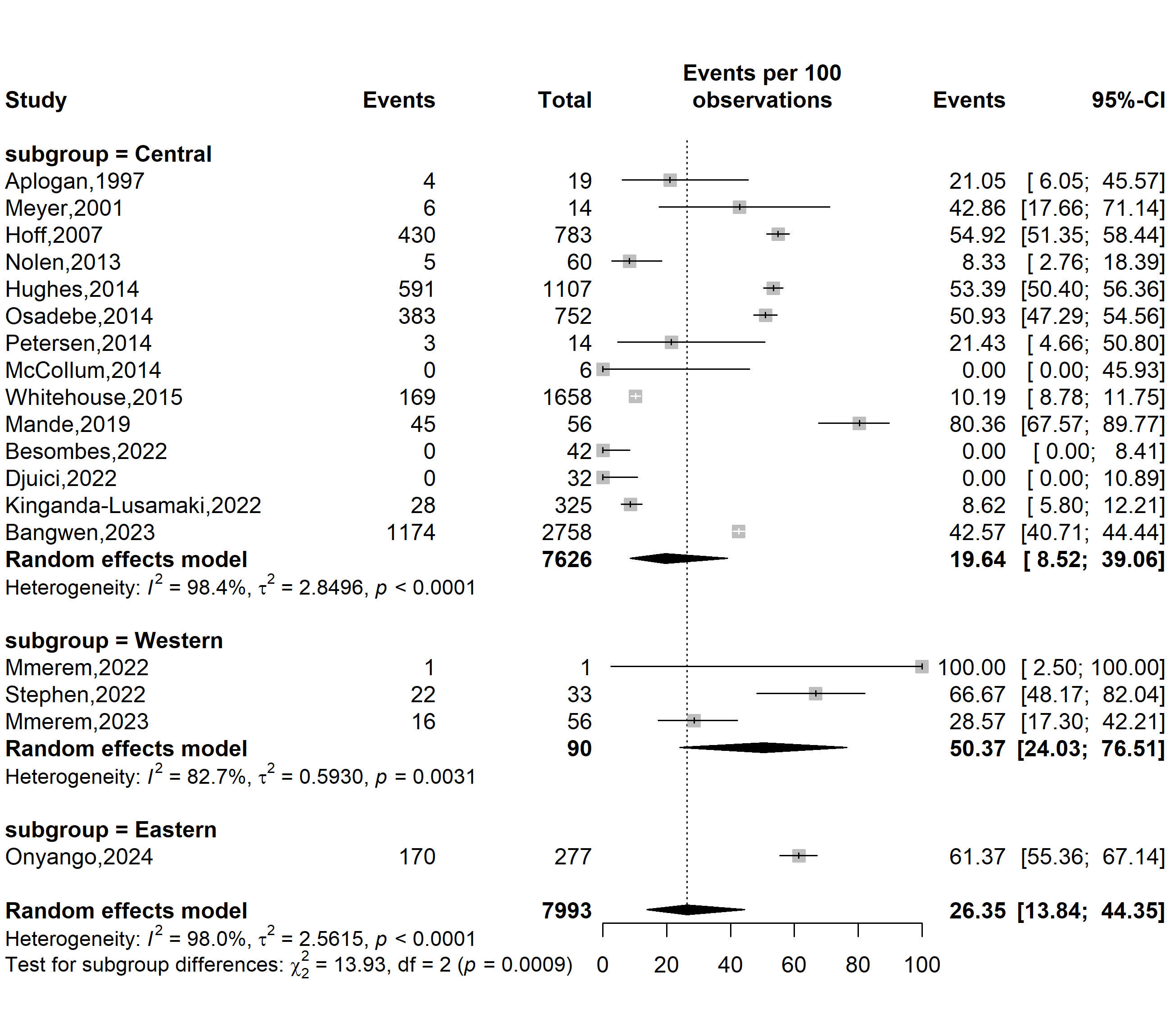

**Supplementary Fig. 6** Prevalence varicella-zoster virus (VZV) coinfections among confirmed mpox cases in Africa by WHO Afro region

**Publication bias**

Egger’s test *p*-value = 0.491

Begg’s test *p*-value = 0.161

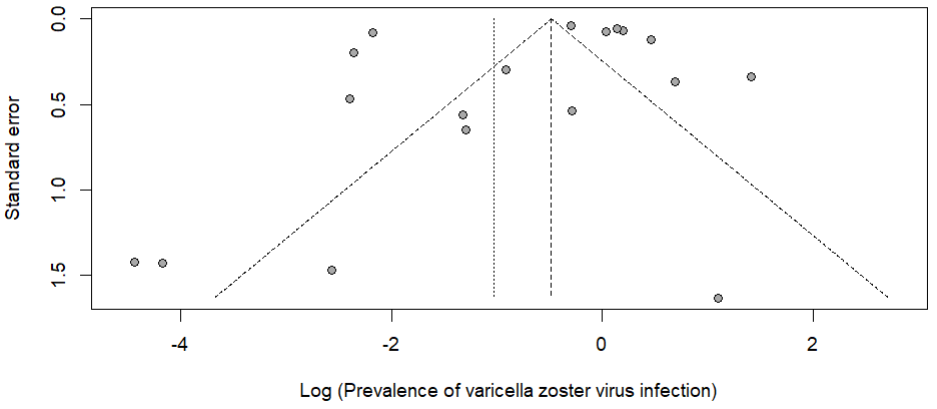

**Supplementary Fig. 7** Funnel plot displaying the pseudo 95% confidence limits and tests assessing the publication bias of studies included

**Sensitivity analysis**

**Event rate (%)**

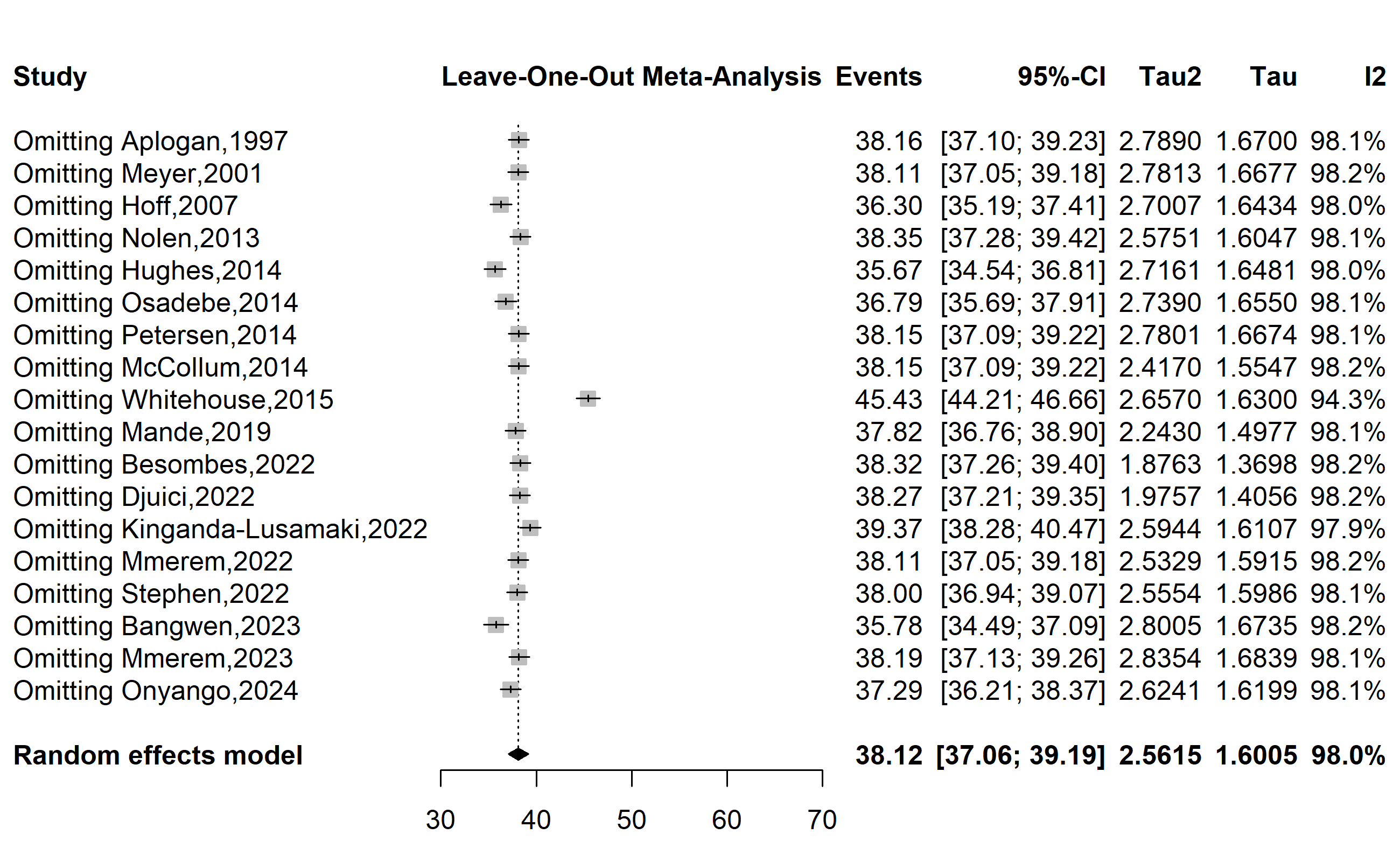

**Supplementary Fig. 8** Sensitivity analysis of the prevalence of varicella-zoster virus coinfections among confirmed mpox cases in Africa
