## Additional Files 4 for "Mpox coinfections and clinical manifestation in Africa: a systematic review and meta-analysis"

**Clade I**

| Clinical manifestation | Frequency (%) |
| --- | --- |
| Rash | 100 |
| Fever | 100 |
| Lymphadenopathy | 100 |
| Sore throat or dysphagia | 100 |
| Myalgia | 80 |
| Oral lesions | 70 |
| Dyspnea | 60 |
| Fatigue | 60 |
| Headache | 50 |
| Cough | 50 |
| Conjunctivitis | 50 |
| Palm lesions | 50 |
| Anorexia | 50 |
| Abdominal pain | 40 |
| Sole lesions | 40 |

|  | Frequency (%) |
| --- | --- |
| Rash | 100 |
| Myalgia | 100 |
| Fatigue | 100 |
| Pruritus or itchy lesion | 93.3 |
| Genital lesions | 92.9 |
| Lymphadenopathy | 90 |
| Palm lesions | 90 |
| Chills or sweat | 90 |
| Headache | 88.9 |
| Sole lesions | 88.9 |
| Light sensitivity | 88.9 |
| Fever | 86.7 |
| Cough | 70 |
| Sore throat or dysphagia | 66.7 |
| Oral lesions | 66.7 |
| Conjunctivitis | 60 |
| Hemorrhagic skin lesions | 22.2 |
| Vomiting or nausea | 20 |
| Bedridden status | 20 |
| Diarrhea | 11.1 |

| Clinical manifestation-Clade I and II | Frequency (%) |
| --- | --- |
| Rash | 96.7 |
| Fever | 84.6 |
| Fatigue | 74.1 |
| Pruritus or itchy lesion | 70.4 |
| Chills or sweat | 66.7 |
| Sore throat or dysphagia | 61.5 |
| Headache | 60 |
| Cough | 48.1 |
| Lymphadenopathy | 46.2 |
| Oral lesions | 42.3 |
| Myalgia | 34.6 |
| Vomiting or nausea | 15.4 |
| Conjunctivitis | 7.7 |

| Clinical manifestation | Frequency (%) |
| --- | --- |
| Rash | 97.6 |
| Pruritus or itchy lesion | 91.5 |
| Genital lesions | 79.4 |
| Painful lesion | 78.2 |
| Fatigue | 75 |
| Malaise | 74.8 |
| Lymphadenopathy | 65.3 |
| Myalgia | 60.6 |
| Anorexia | 56.9 |
| Headache | 51 |
| Sore throat or dysphagia | 49.1 |
| Dysuria | 39.5 |
| Palm lesions | 38.7 |
| Cough | 31.6 |
| Anal lesions | 31.6 |
| Abdominal pain | 30.2 |
| Sole lesions | 23.2 |
| Oral lesions | 22.9 |
| Vomiting or nausea | 22.7 |
| Painful eye | 20 |
| Fever | 16.8 |
| Dyspnea | 13.8 |
| Diarrhea | 12.6 |
| Rectal pain | 10.8 |
| Visual problem | 9.9 |
| Bedridden status | 9.3 |
| Chills or sweat | 11.1 |
| Hematuria | 5.1 |
| Conjunctivitis | 3.8 |
| Confusion | 2.8 |
| Convulsions | 0.2 |

|  | Frequency (%) |
| --- | --- |
| Rash | 100 |
| Fever | 88.5 |
| Headache | 74.5 |
| Pruritus or itchy lesion | 71.7 |
| Palm lesions | 68.6 |
| Lymphadenopathy | 67.4 |
| Genital lesions | 66.3 |
| Sole lesions | 63.6 |
| Chills or sweat | 62.6 |
| Malaise | 61.9 |
| Sore throat or dysphagia | 58.4 |
| Myalgia | 53.4 |
| Fatigue | 51.7 |
| Joint pain | 23.8 |
| Oral lesions | 32.4 |
| Cough | 26.6 |
| Conjunctivitis | 22.3 |
| Vomiting or nausea | 20.9 |
| Light sensitivity | 20.9 |
| Anorexia | 9.5 |
| Dehydration | 9.5 |
| Hepatomegaly | 9.5 |
| Diarrhea | 4.8 |
| Scrotal edema | 4.8 |

**Fig. 1** Clinical manifestations of the disease in Africa by mpox viral strains
